# Mucosal versus systemic antibody responses to SARS-CoV-2 antigens in COVID-19 patients

**DOI:** 10.1101/2020.08.01.20166553

**Authors:** Baweleta Isho, Kento T. Abe, Michelle Zuo, Alainna J. Jamal, Bhavisha Rathod, Jenny H. Wang, Zhijie Li, Gary Chao, Olga L. Rojas, Yeo Myong Bang, Annie Pu, Natasha Christie-Holmes, Christian Gervais, Derek Ceccarelli, Payman Samavarchi-Tehrani, Furkan Guvenc, Patrick Budylowski, Angel Li, Aimee Paterson, Yue Feng Yun, Lina M. Marin, Lauren Caldwell, Jeffrey L. Wrana, Karen Colwill, Frank Sicheri, Samira Mubareka, Scott D. Gray-Owen, Steven J. Drews, Walter L. Siqueira, Miriam Barrios-Rodiles, Mario Ostrowski, James M. Rini, Yves Durocher, Allison J. McGeer, Jennifer L. Gommerman, Anne-Claude Gingras

**Affiliations:** Department of Immunology, University of Toronto, Toronto, ON, Canada; Lunenfeld-Tanenbaum Research Institute at Mount Sinai Hospital, Sinai Health System, Toronto, ON, Canada; Department of Molecular Genetics, University of Toronto, Toronto, ON, Canada; Institute of Health Policy, Management and Evaluation, University of Toronto, Toronto, ON, Canada; Department of Microbiology, at Mount Sinai Hospital, Sinai Health System, Toronto, ON, Canada; Combined Containment Level 3 Unit, University of Toronto, Toronto, ON, Canada; Mammalian Cell Expression, Human Health Therapeutics Research Centre, National Research Council Canada, Montréal, QC, Canada; Institute of Medical Science, University of Toronto, Toronto, ON, Canada; College of Dentistry, University of Saskatchewan, Saskatoon, SK, Canada; Department of Laboratory Medicine and Molecular Diagnostics, Division of Microbiology, Sunnybrook Health Sciences Centre; Biological Sciences, Sunnybrook Research Institute; and Division of Infectious Diseases, Sunnybrook Health Sciences Centre, Toronto, ON, Canada; Department of Laboratory Medicine and Pathology, University of Toronto, Toronto, ON, Canada; Canadian Blood Services, Edmonton, AB & Department of Laboratory Medicine and Pathology, University of Alberta, Edmonton, AB, Canada; St. Michael’s Hospital, Toronto, ON, Canada; Li Ka Shing Knowledge Institute; Department of Medicine, University of Toronto, Toronto, ON, Canada; Department of Biochemistry, University of Toronto, Toronto, ON, Canada

## Abstract

While the antibody response to SARS-CoV-2 has been extensively studied in blood, relatively little is known about the mucosal immune response and its relationship to systemic antibody levels. Since SARS-CoV-2 initially replicates in the upper airway, the antibody response in the oral cavity is likely an important parameter that influences the course of infection, but how it correlates to the antibody response in serum is not known. Here, we profile by enzyme linked immunosorbent assays (ELISAs) IgG, IgA and IgM responses to the SARS-CoV-2 spike protein (full length trimer) and its receptor binding domain (RBD) in serum (n=496) and saliva (n=90) of acute and convalescent patients with laboratory-diagnosed COVID-19 ranging from 3–115 days post-symptom onset (PSO), compared to negative controls. Anti-CoV-2 antibody responses were readily detected in serum and saliva, with peak IgG levels attained by 16–30 days PSO. Whereas anti-CoV-2 IgA and IgM antibodies rapidly decayed, IgG antibodies remained relatively stable up to 105 days PSO in both biofluids. In a surrogate neutralization ELISA (snELISA), neutralization activity peaks by 31–45 days PSO and slowly declines, though a clear drop is detected at the last blood draw (105–115 days PSO). Lastly, IgG, IgM and to a lesser extent IgA responses to spike and RBD in the serum positively correlated with matched saliva samples. This study confirms that systemic and mucosal humoral IgG antibodies are maintained in the majority of COVID-19 patients for at least 3 months PSO. Based on their correlation with each other, IgG responses in saliva may serve as a surrogate measure of systemic immunity.

**One Sentence Summary:** In this manuscript, we report evidence for sustained SARS-CoV-2-specific IgG and transient IgA and IgM responses both at the site of infection (mucosae) and systemically in COVID-19 patients over 3 months and suggest that saliva could be used as an alternative biofluid for monitoring IgG to SARS-CoV-2 spike and RBD antigens.

## Introduction

Antibodies play an important role in neutralizing virus and provide protection to the host against viral re-infection. The antibody response to SARS-CoV-2 infection has been extensively studied in the blood (serum, plasma) of COVID-19 patients in order to gain insights into the host immune response. Antibody levels to the spike protein are particularly important since this large trimeric glycoprotein harbours the receptor binding domain (RBD). The RBD facilitates SARS-CoV-2 access to human cells by binding to its counter receptor angiotensin converting enzyme 2 (ACE-2) (*1*), and neutralizing antibodies have been shown to target the RBD (*2*). Most studies agree that the IgG antibodies to SARS-CoV-2 spike and RBD antigens are detected in the blood of greater than 90% of subjects by 10–11 days post-symptom onset (PSO) (*3-7*). However, whether levels of IgG specific for SARS-CoV-2 antigen persist (*8, 9*), or alternatively decay (*10*), remains a debated issue. Examination of different biofluids from multiple cohorts, and attention to the antigens tested, is required to resolve this extremely important issue that has high relevance to vaccine design.

Another gap in our knowledge is that we know very little about the local antibody response at the site of infection. SARS-CoV-2 enters the naso- and oro-pharyngeal tracts where it subsequently replicates (*11*). For this reason, nasopharyngeal and throat swabs are used to test for virus using reverse transcriptase quantitative PCR (RT-qPCR) to detect viral RNA. However, saliva has also been shown to be an effective biofluid for testing for the presence of SARS-CoV-2 mRNA (*12-15*). This makes sense given that pharyngeal SARS-CoV-2 shedding precedes viral replication in the lungs (*11*), and, like cytomegalovirus (*16, 17*), the salivary glands themselves can be a reservoir for the virus (*18*). Yet in spite of the oral cavity being a site for viral replication, few studies have examined anti-SARS-CoV-2 antibodies in this compartment.

In this study, we examined the anti-SARS-CoV-2 antibody response over a 115-day period in the serum and saliva of n=496 (serum) and n=90 (saliva) samples from patients with COVID-19, compared to controls. Antigen-specific IgG in both biofluids were maximally detected by 16–30 days PSO and did not drastically decline in their relative levels as late as 100–115 days PSO. In contrast, antigen-specific IgM and IgA were rapidly induced but subsequently declined in both serum and saliva. In serum, neutralizing antibodies reached their maximum by 31–45 days PSO and slowly declined up to 105 days, with a more pronounced drop in the last blood draw (105–115 days PSO) Importantly, IgG and IgM levels against both antigens were strongly correlated across paired serum and saliva samples (n=71), indicating that saliva can be used for monitoring the immune response to SARS-CoV-2 infection. Taken together, the systemic and mucosal IgG response to SARS-CoV-2 is sustained over a 3-month period, while the IgM and IgA response occurs early and is transient.

## Results

*A chemiluminescent fully automated method for detecting antibodies to SARS-CoV-2 antigens in the serum of acute and convalescent patients*. To study the antibody response to SARS-CoV-2, we initially focused on antibodies (IgM, IgG, IgA) to the spike homotrimer and the RBD, since neutralizing antibodies are directed to the spike protein (*19*). Enzyme-linked immunosorbent assays (ELISAs) for the detection in serum (or plasma) of anti-spike trimer and anti-spike RBD antibodies were built as in (*3, 20*) as 96-well colorimetric assays, and implemented as automated 384-well chemiluminescence assays (see Methods). Receiver-Operating Characteristic (ROC) curves were generated on cohorts of true negatives (banked samples collected pre-COVID, n=339 for manual and automated assays) and positives (convalescent patients with confirmed PCR diagnostic, n=402 for manual and automated assays, see Table 1). For manual and automated IgG assays, sensitivities of 95.6% and 95.5% for spike and 93.8% and 91.3% for RBD, respectively, at a false positive rate of ≤1%, were obtained in these cohorts (Supplemental Figure 1A-B, and Supplemental Table 1 for ROC statistics). The Areas Under the Curves (AUCs) were ≥0.97 in all cases, indicating excellent assay performance. Automated assays for the detection of IgA and IgM were also developed (Supplemental Figure 1C-D). The results for the automated and manual IgG assays were well correlated (Supplemental Figure 1E-F).

**Table 1.** Cohorts of patients and negative controls.

|  |  | SALIVA |  |  |  |  |  | BLOOD |  |  |  |  |
| --- | --- | --- | --- | --- | --- | --- | --- | --- | --- | --- | --- | --- |
|  |  | No. patient | No. samples | Median Age | Sex |  |  | No. patients | No. samples | Median Age | Sex |  |
|  |  |  |  |  | No. M | No. F |  |  |  |  | No. M | No. F |
| **All samples** |  | 247 | 263 | - | 141 | 106 |  | 777 | 835 |  |  |  |
| Patients with COVID-19 | Cohort 1 | 47 | 54 | 61 | 28 | 19 | Patients with COVID-19 | 438 | 496 | 58 | 262 | 234 |
|  | Cohort 2 | 81 | 90 | 58 | 48 | 33 | Pre-COVID Negative Controls | 300 | 300 | 54.5 | 150 | 150 |
| Pre-COVID Negative Controls | Cohort 1 | - | - | - | - | - |  |  |  |  |  |  |
|  | Cohort 2 | 27 | 27 | 43 | 12 | 15 |  |  |  |  |  |  |
| Unexposed Negative Controls Collected in 2020 | Cohort 1 | 42 | 42 | 60 | 24 | 18 |  |  |  |  |  |  |
|  | Cohort 2 | 50 | 50 | 58 | 29 | 21 |  |  |  |  |  |  |
| *Matched saliva-serum samples* |  | 71 | 71 | 58 | 33 | 38 |  |  |  |  |  |  |

These automated ELISA assays were used to profile cohorts of confirmed acute and convalescent sera from COVID-19 patients collected as part of COVID-19 surveillance by the Toronto Invasive Bacterial Diseases Network (Table 1). As expected, based on the ROC analysis, the convalescent and pre-COVID controls had very different ratio distributions for both antigens (Figure 1A, D). On the other hand, serum collected from patients less than 21 days PSO (acute serum, n=132) had bimodal distributions in their IgG responses for both antigens (with an overall lower mean), suggesting that antibody concentrations were increasing over time. To compare the relationship between RBD and spike trimer IgG levels, we plotted their values against each other. While there was an overall high correlation between the antigens (Figure 1G), we noted many more acute specimens with high spike-trimer and low RBD response than vice versa, consistent with the fact that RBD is included within the spike trimer antigen. The concentration of IgA and IgM in convalescent serum was also clearly higher than that of the pre-COVID samples, but the acute cases had a higher median than the convalescents (Figure 1B & E, C & F). The IgA and IgM levels to RBD and spike were also well correlated (Figure 1H-I).

**Figure 1.**
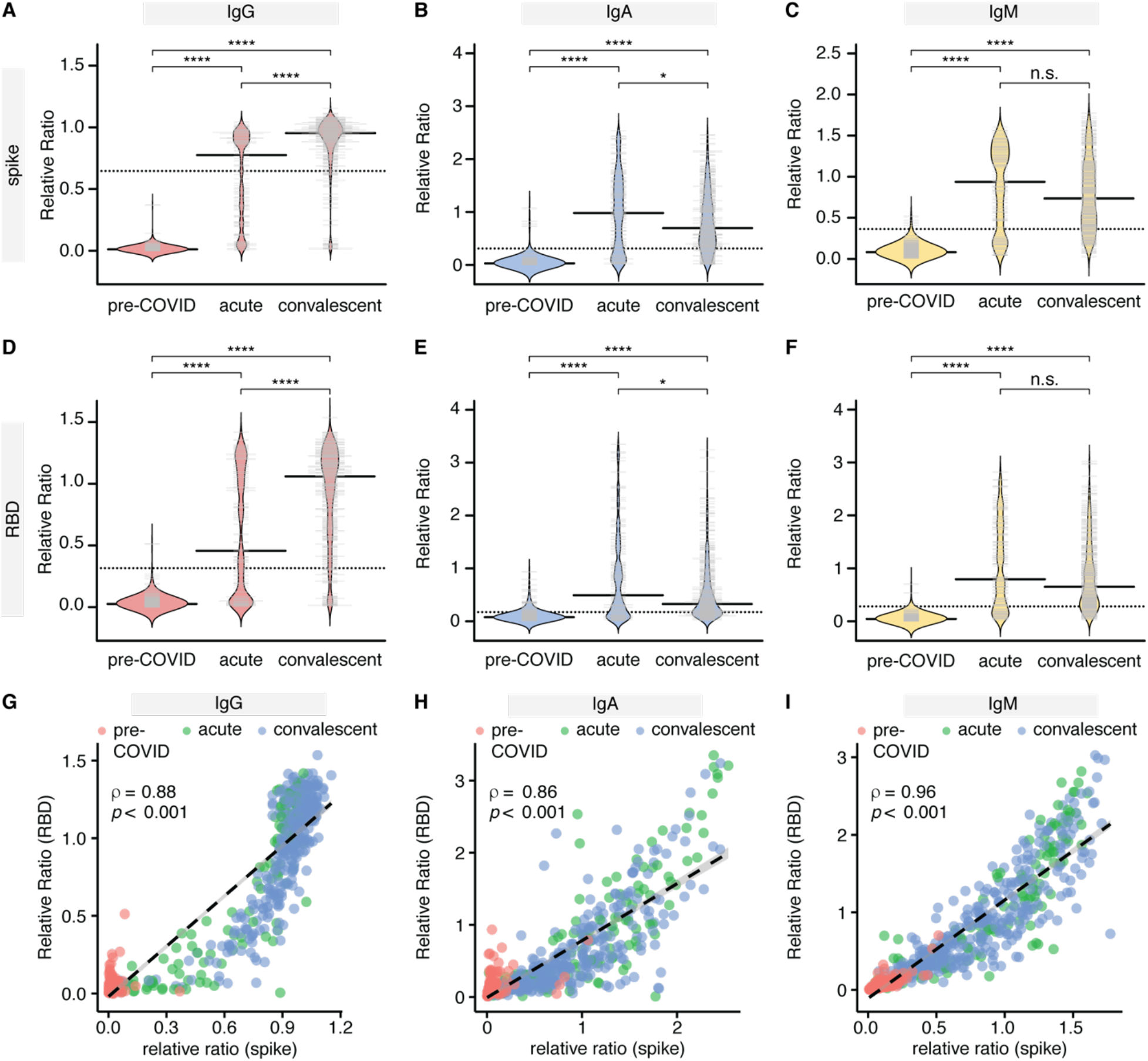
Cross-sectional analysis of IgG and IgA responses to the spike and RBD antigens of SARS-CoV2 in serum. (**A-F**) Indicated immunoglobulins to spike and RBD were profiled by ELISA in cohorts of pre-COVID samples (n=300), hospitalized patients with acute COVID infection (n=139) and convalescent patients (n=357). All data, expressed as ratios to a pool of convalescent samples, were plotted using bean plots. Solid bars denote the median and dotted line represents the median across all samples used in the plot. (**G-I**) levels of IgG (G), IgA (H) and IgM (I) to the RBD (y-axis) and spike (x-axis) antigens for the indicated patient groups. Spearman correlation coefficient is indicated. Mann-Whitney U test for significance was performed. n.s = not significant, *= p ≤ 0.05, **** = p < 0.0001.

The bimodal distribution of the IgG responses in the acute serum (Figure 1A, D), along with the different patterns of response for IgG versus IgA/IgM in acute and convalescent specimens (Figure 1B & E, C & F), prompted us to plot the antibody levels against days PSO. Spearman rank correlation analysis revealed an overall increase in the IgG response versus a decrease in the IgA and IgM response to both antigens over time, and the IgG response in particular did not appear to be linear (compare panels A-B to C-D and E-F in Supplemental Figure 2; IgG results were reproduced in the analysis of the manual IgG assays, shown in panels G-H). To look at this response more closely, specimens were binned by days PSO (15-day intervals), and the levels of the different immunoglobulins were plotted (the pre-COVID negative control samples were plotted alongside for comparison; Figure 2). As was reported in other studies (*3*, *4, 7*), the IgG levels reached peak in the 16–30 days bin, and the levels of IgG against spike trimer were sustained over 115 days (<7.3% change in the median as compared to the maximum; Figure 2A). However, IgG levels against RBD showed a ~25.3 % decrease by day 105, and ~46.0 % by day 115 (Figure 2D). The behavior of IgA and IgM to both antigens was by contrast much less sustained: after reaching a maximum in the 16–30 days bin, there was a clear and continuous decline throughout the time series such that by 115 days, the anti-spike and anti-RBD IgA levels were ~74.1 % and ~84.2 % of their respective maximal levels, while IgM levels were ~66.2 % and ~75.1 %, respectively (Figure 2B, E & C, F). Multivariable analyses adjusting for severity of illness, sex, and patient age, did not change conclusions about the aforementioned relationships between time PSO and anti-RBD IgM, anti-spike IgM, anti-RBD IgA, anti-spike IgA, and anti-RBD IgG; however, the modest decline in anti-spike IgG after day 35 was statistically significant (data not shown).

**Figure 2.**
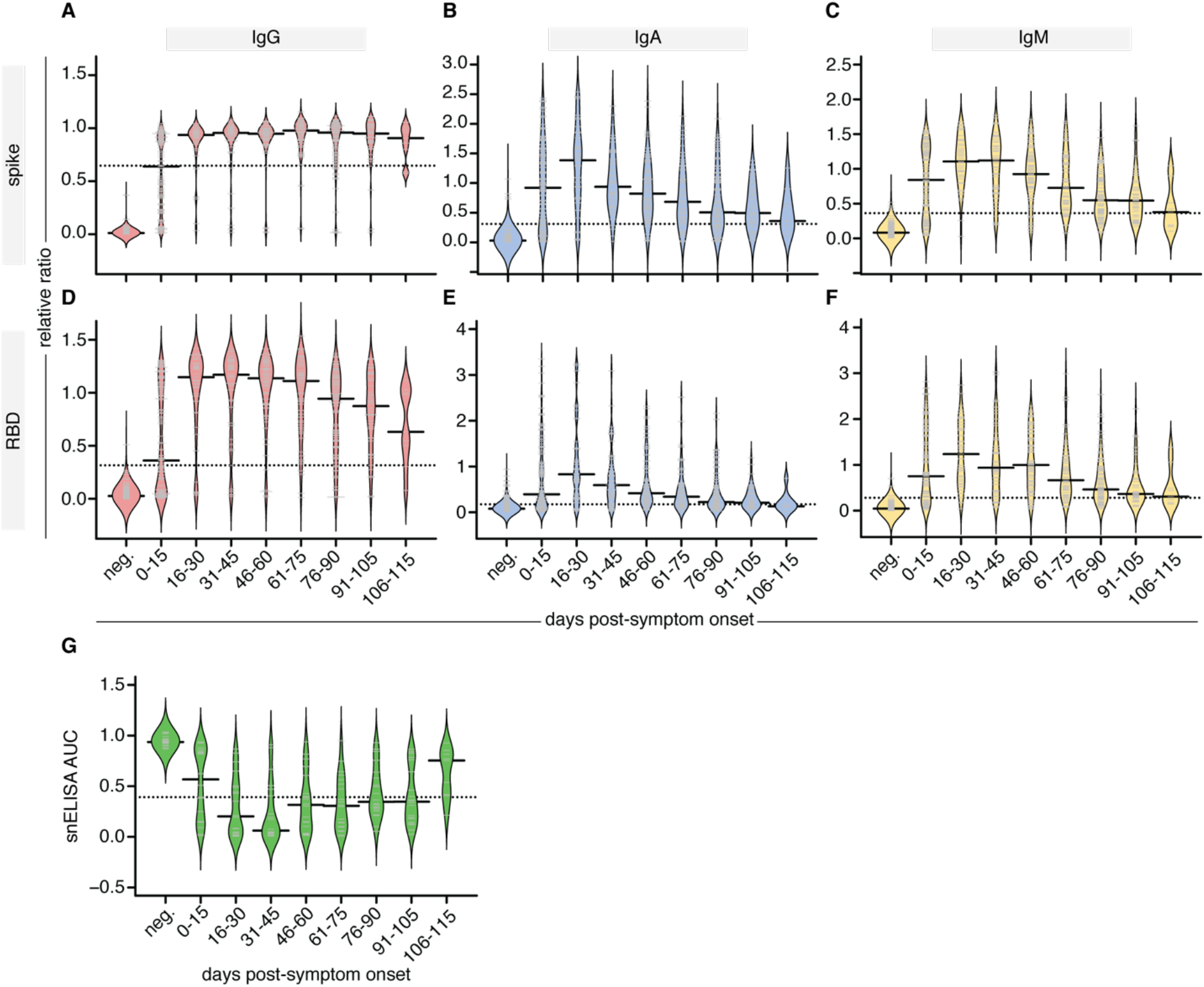
Persistence of antibodies in the serum of affected individuals. (**A-F**) Binned relative ratios (to a pool of positive controls) of spike (**A-C**) and RBD (**D-F**) to the indicated antibodies, displayed as bean plots. (**G**) The AUC results of the surrogate neutralization ELISA are also shown. Days PSO are binned in 15-day increments and are compared to pre-COVID samples (neg). Solid bars denote the median and dotted line represents the median across all samples used in the plot.

To dissect these results, we analyzed pairs of serum samples from the hospitalized patients (n=58), collected at admission, and 3–12 weeks later, and performed a non-parametric loess analysis (as in (*21*)). These results depict a relative stability of the IgG anti-spike trimer levels, a partial decrease in the anti-RBD IgG and anti-spike IgA levels, and a near complete loss in the anti-RBD IgM and IgA levels over time (Figure 3).

**Figure 3.**
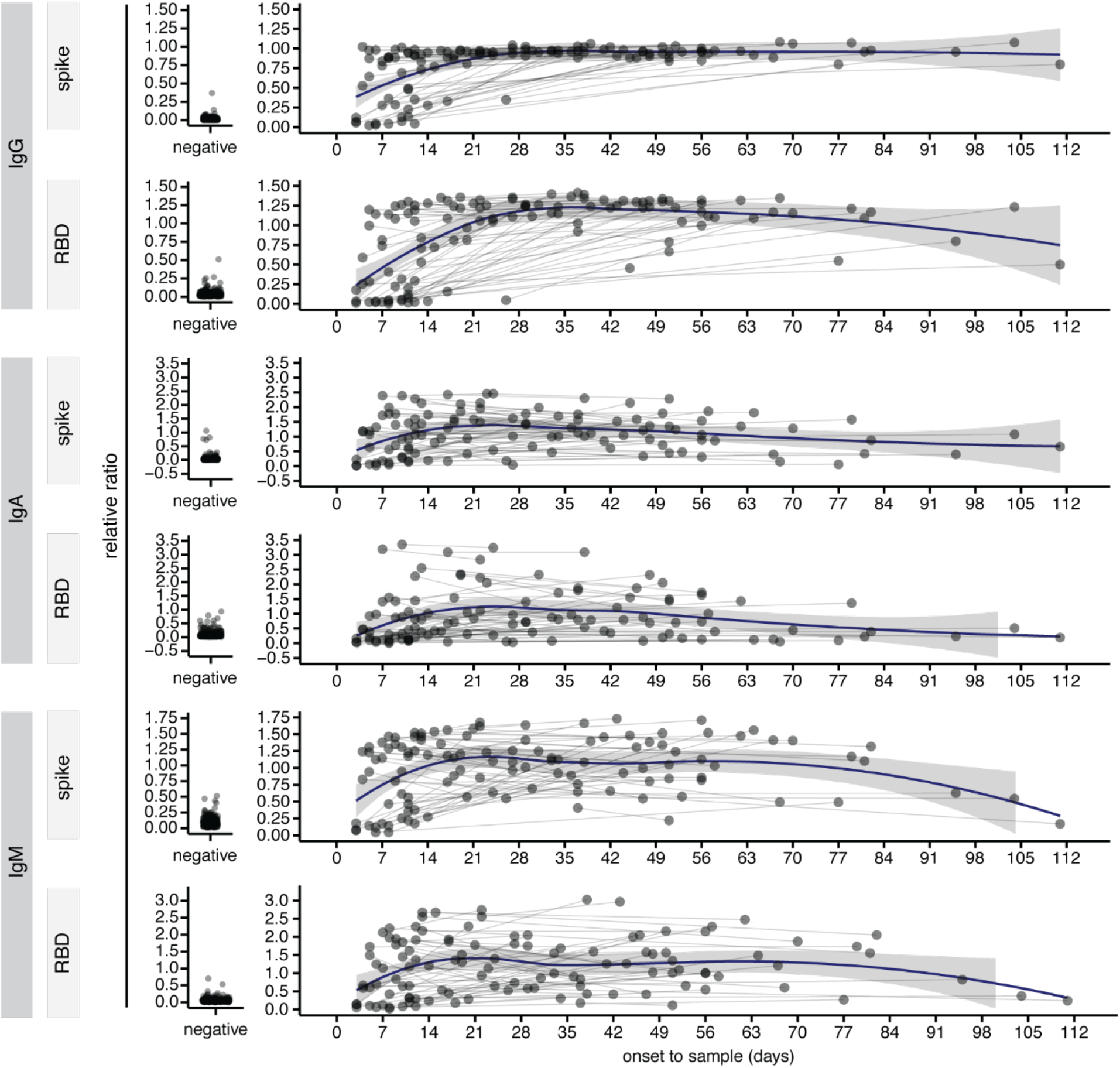
A longitudinal analysis of IgG and IgA responses to the spike and RBD antigens of SARS-CoV2 in serum. Analysis of the changes in the indicated Ig-antigen levels in patients profiled twice, in comparisons to the relative levels in pre-COVID negative controls (left). Dots represent individual serum samples collected at the indicated times, and the samples from the same patients are connected by the lines. A non-parametric loess function is shown as the blue line, with the grey shade representing the 95% confidence interval.

Although our focus was on the spike protein, we also examined the antibody response to nucleocapsid (a.k.a. nucleoprotein, NP), since this is the antigen targeted by multiple commercial assays. We developed an assay using bacterially-expressed NP (Supplemental Figure 3A-C). When we examined the levels of anti-NP antibodies binned for time PSO, we found that their patterns closely resembled those for anti-spike and anti-RBD IgG and IgA/IgM responses, namely a relative stability in the IgG and more rapid decline in IgA/IgM levels in both the binned time series and the longitudinal series (Supplemental Figure 3D-F).

To evaluate the neutralization potential of these antibodies, we used our recently established protein-based surrogate neutralization ELISA (snELISA) approach (Figure 2G;(*20*)). Briefly, the snELISA measures the ability of antibodies (in serum in our case) to prevent the association of soluble biotinylated ACE2 to immobilized RBD: a higher signal (snELISA AUC) in this assay indicates low neutralization. Using the binned time series as above, we report that the neutralization reaches its maximum in the 31–45 day PSO bin, and decreases to an intermediate median plateau in the 46–105 day PSO bins before more drastically dropping in the 106–115 day PSO samples (we note, however, that fewer samples are in this time bin (n=9) compared to the other bins (n=20); Figure 2G).

In summary, in a large cross-sectional survey, IgG, but not IgA or IgM levels persisted for at least 3 months PSO for all antigens measured, with the levels of antibodies to the spike trimer being more stable over time than those to the RBD and NP. Neutralizing antibodies levels mirrored these antibody levels, though the drop observed in the last bin (105–115 days PSO), which was not as powered as the other bins, will need to be investigated more closely.

*Antibodies to SARS-CoV-2 antigens are detected in the saliva of COVID-19 patients*. While our serum-based assays are scalable and robust, saliva represents a relatively unexplored biofluid for detecting antibodies to SARS-CoV-2 antigens with many practical benefits, including being non-invasive and the capacity for self-collection at home. The disadvantage of saliva as a biofluid is its very low concentration of antibodies (*22*), making it necessary to optimize the sensitivity of detection. We explored various detection methods and found that plating biotinylated antigen onto streptavidin coated plates was required to obtain reliable signal-to-noise ratios; the method also required that the saliva be pre-adsorbed to remove any streptavidin-binding protein (data not shown). While heat (65°C for 30 min) prevented detection of antibodies in the saliva, incubation with Triton X-100 was compatible with our assay (Supplemental Figure 4) and resulted in viral inactivation (Supplemental Table 2). Bolstered by these findings, we first performed a pilot experiment, using expectorated saliva samples acquired during the early phase of the pandemic, measuring antibody levels to SARS-CoV2 antigens in n=54 COVID-19 patients (cohort 1), comparing to unexposed negative controls collected locally (n=42). Since these samples were diluted to varying degrees, we normalized values to total IgG/IgA (depending on the isotype assay) or to albumin levels as done before by others (*23*). Saliva samples from COVID-19 patients displayed a significantly higher level of IgG and IgA levels to spike and RBD compared to negative controls when normalized with either method (Supplemental Figure 5).

Following this pilot experiment, we proceeded with further saliva collections using a standardized collection method without a diluent (cohort 2) in order to measure IgG, IgA and IgM levels to both spike and RBD antigens. In cohort 2, we obtained n=90 samples from 80 patients ranging in time PSO from day 3–104. These were compared to 50 unexposed negative controls for cohort 2, of which 42 were also negative controls for cohort 1. To these negative controls, we also added pre-COVID era saliva samples as an additional comparator (n=27). We performed a normalization to internal plate controls (pooled saliva from several COVID-19 patients) as shown in Supplemental Figure 6. Total IgG levels, but not IgA or IgM levels, were found to be higher in COVID-19 patients compared to controls (Figure 4A-C). Moreover, cohort 2 exhibited statistically significant differences between the relative levels of IgG, IgA and IgM antibodies specific to spike and RBD antigens compared to saliva from negative controls (Figure 4D-I). The sensitivity of the saliva assays for IgG antibodies to spike and RBD (at a false discovery rate <2%) were 89% and 85%, respectively, while the sensitivity of the assays for IgA antibodies to spike and RBD were 51% and 30%, respectively, and the sensitivity of the assays for IgM antibodies to spike and RBD were 57% and 33%, respectively. (Supplemental Figure 7 and Supplemental Table 3). The lower sensitivity of the IgA assays is attributed in part to the higher levels of anti-spike and anti-RBD IgA levels in the negative controls (see Discussion).

**Figure 4.**
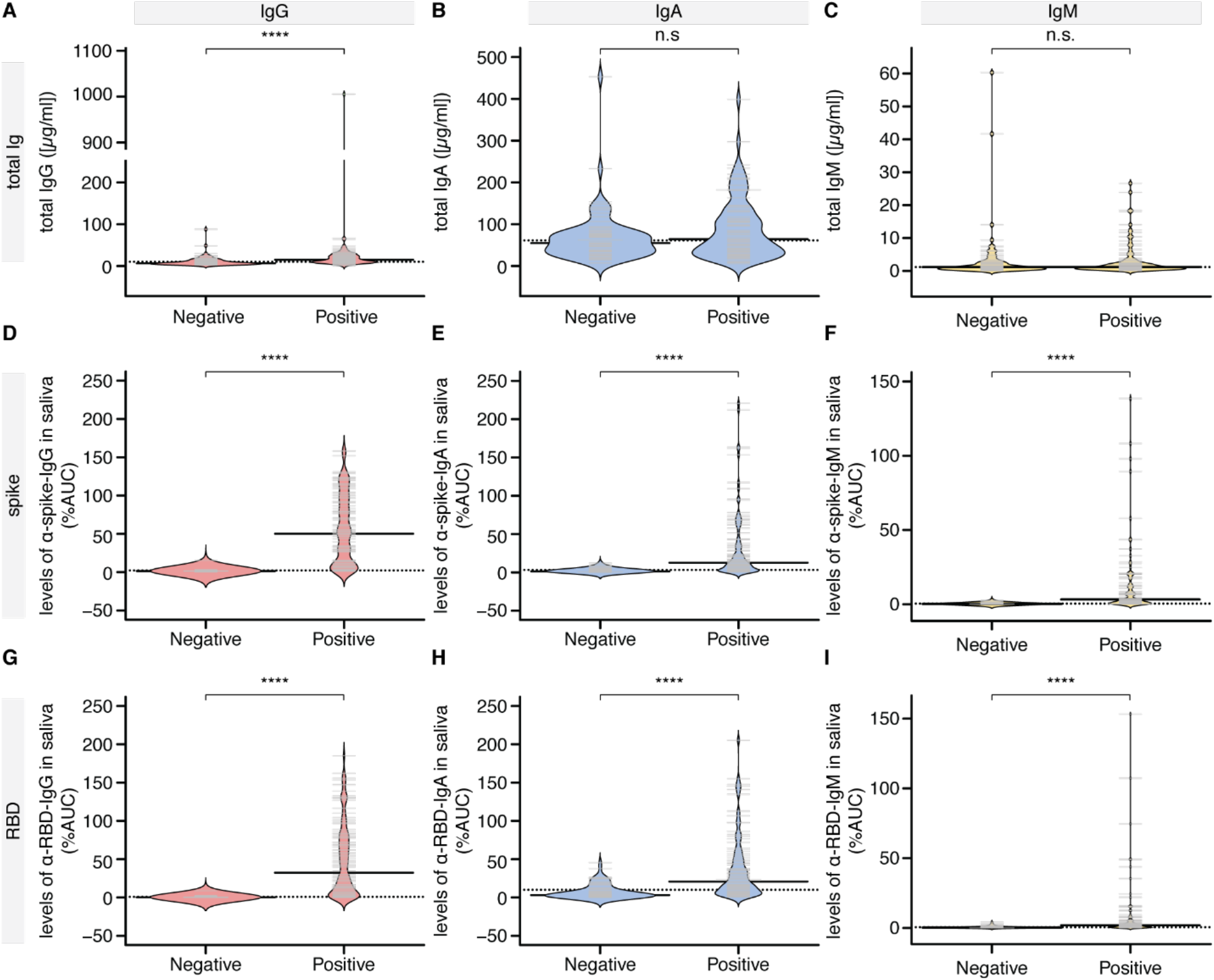
Cross-sectional analysis of antibody responses to the spike and RBD antigens of SARS-CoV-2 in saliva. Saliva specimens from the cohort of COVID-19 patients were tested for the presence of IgG, IgA and IgM antibodies to SARS-CoV-2 spike and RBD antigens (Positive), comparing with a mixture of unexposed asymptomatic controls collected locally and pre-COVID era controls (Negative). In these cohort 2 samples collected in Salivettes® we had sufficient material to perform several dilutions and to generate an AUC value for each subject. This value was expressed as a percentage of the AUC derived from a control sample consisting of pooled saliva from COVID-19 patients. Because the saliva was not diluted during collection, we were able to derive the concentration of antibodies in both negative controls and COVID-19 patients. (**A-C**) Total IgG, IgA and IgM levels in the saliva. (**D-I**) Saliva data for negative controls versus COVID-19 patients. Solid bars denote the median and dotted line represents the median across all samples used in the plot. Mann-Whitney U test for significance was performed. **** = p < 0.0001, n.s. = not significant.

Next, we examined the levels of anti-spike and anti-RBD antibodies in our cross-sectional cohort over time PSO. Similar to the serum data, IgG levels in saliva to the spike and RBD antigens remained stable throughout the 3-month collection period. In contrast, significant decreases were observed for IgA levels to spike and RBD (ρ=-0.307 and ρ=-0.300, respectively), and similar results were observed for IgM levels to spike and RBD (ρ=-0.33 and ρ=-0.32, respectively). By day 100, anti-spike and anti-RBD IgA and IgM levels were barely detectable (Figure 5). In summary, infection with SARS-CoV-2 results in detectable IgG, IgA and IgM response in saliva against the spike and RBD antigens, with only the IgG response persisting beyond day 60.

**Figure 5.**
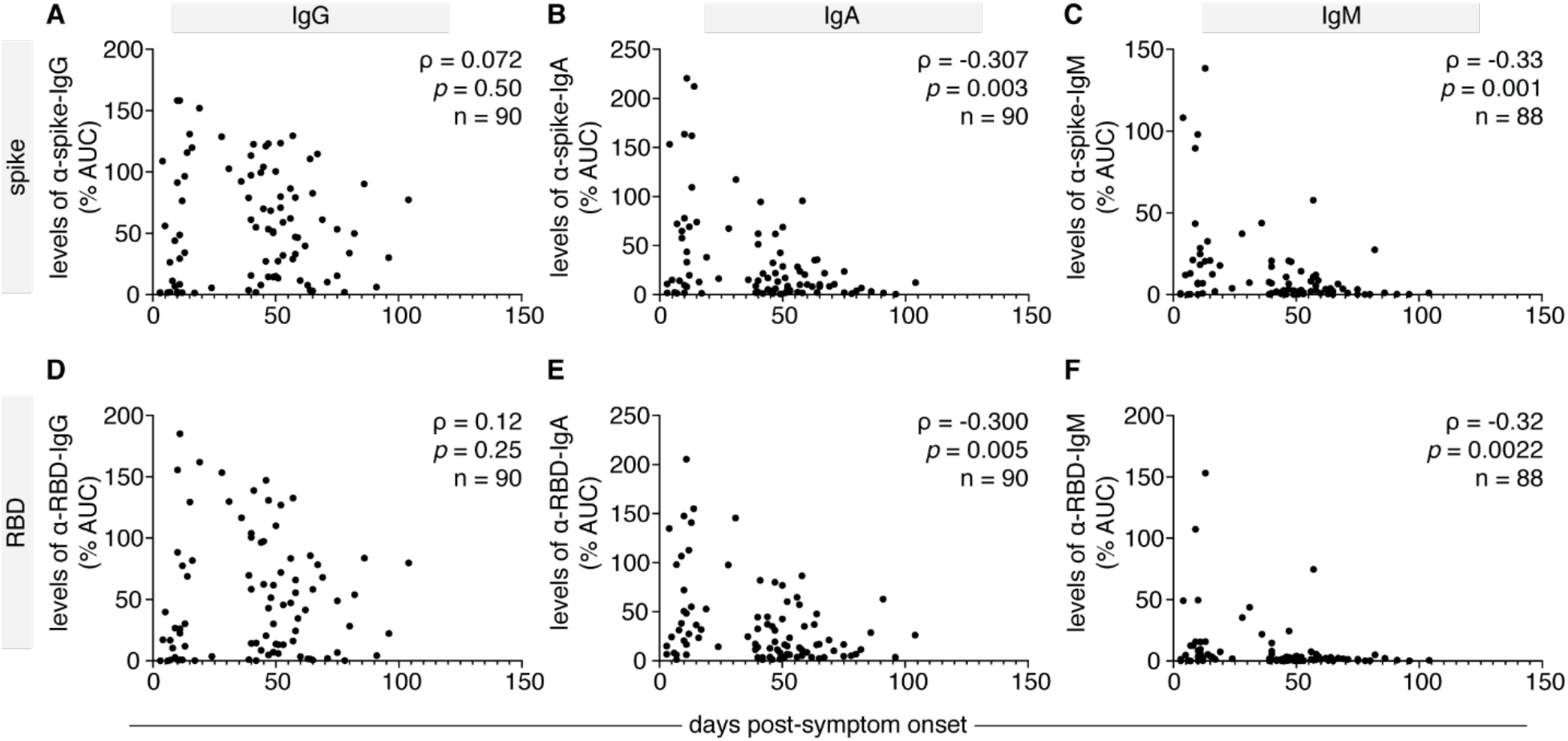
A cross-sectional analysis of antibody responses to the spike and RBD antigens of SARS-CoV-2 in saliva correlated with time PSO. A second cohort of COVID-19 patients (n=90) was tested for the presence of IgG and IgA antibodies to SARS-CoV-2 spike and RBD antigens in the saliva, comparing with a mixture of unexposed negative controls collected locally and pre-COVID era negative controls. (**A-F**) Saliva data for all 6 antigen-specific ELISA readouts plotted as time PSO. Spearman correlation coefficients (ρ) and p-value are indicated. In multivariable analysis adjusted for age, sex and severity of illness, there was a significant decline in anti-RBD and anti-spike IgA, but not significant change in the level of anti-RBD or anti-spike IgG.

*Antibody levels to SARS-CoV-2 antigens in the serum correlate with those in the saliva*. As mentioned, saliva has many advantages for biofluid collection over serum. To assess whether saliva might be reliably used in a diagnostic test, we determined whether the antibody levels to spike and RBD in the saliva correlated with those measured in the serum (Figure 6). Of the COVID-19 patients analyzed, n=71 had paired saliva and serum samples taken at similar timepoints (i.e. within 4 days). When comparing the saliva %AUC values to the ratios in serum, we observed a significant positive correlation between saliva and serum for each antigen-antibody combination. Correlations for anti-RBD and anti-spike IgG (ρ=0.71, ρ=0.54), and anti-RBD and anti-spike IgM (ρ=0.65, ρ=0.58) were all reasonably strong. The correlations between the levels of serum and saliva anti-RBD and anti-spike IgA were more modest (ρ=0.39 and 0.54 respectively). Therefore, at least for anti-spike IgM and anti-RBD IgG measurements, saliva may represent a good alternative for antibody testing.

**Figure 6.**
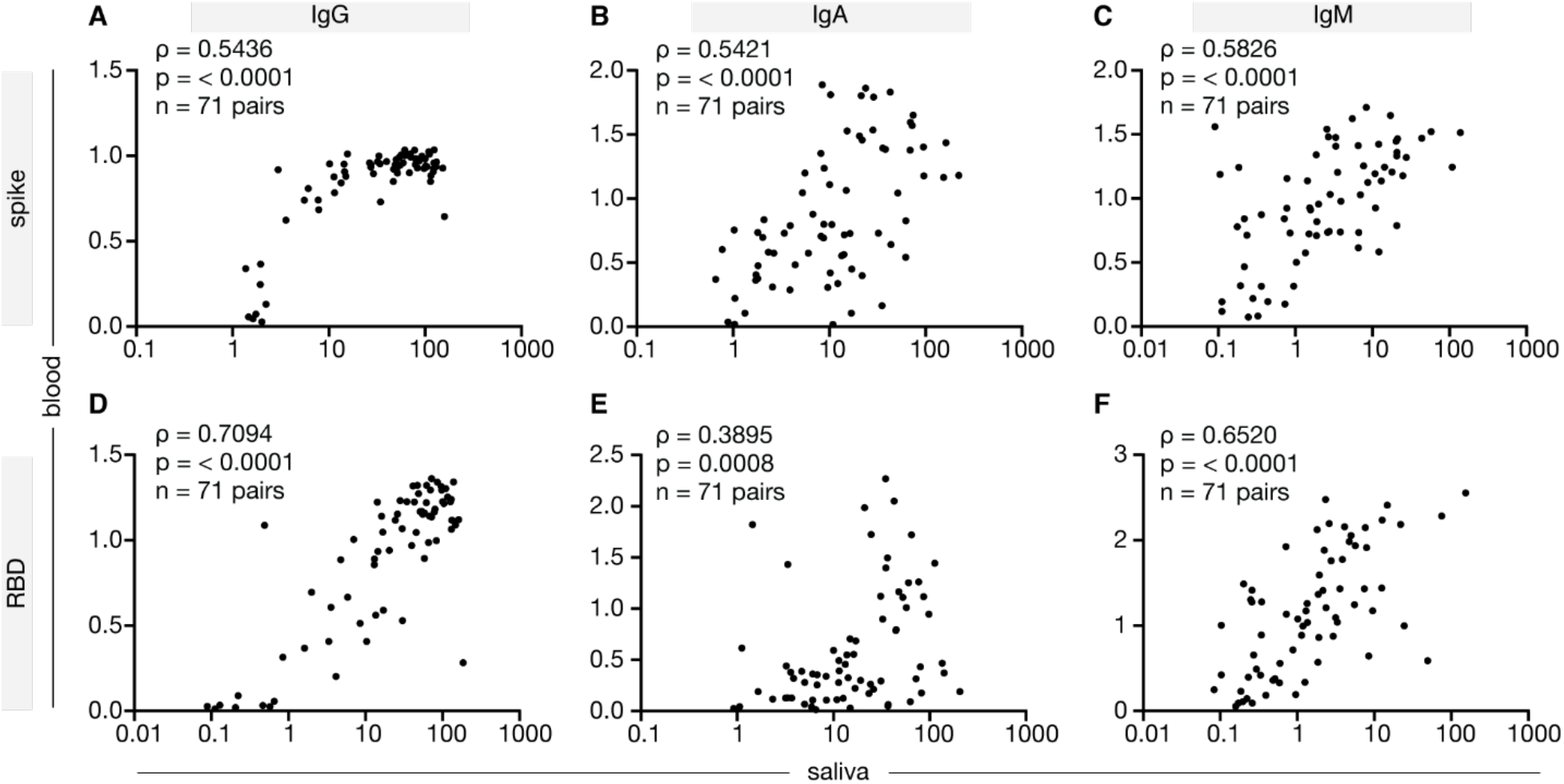
Correlation of IgG, IgA and IgM responses to the spike and RBD antigens in serum and saliva. (**A-F**) A subset of blood and saliva sample pairs (n=71) collected from the same patient within 4 days were analyzed for correlations in levels of anti-spike and anti-RBD IgG, IgA and IgM antibodies. Spearman correlation coefficient (ρ) and p-value are indicated.

## Discussion

Antibodies are key components in the arsenal of protective immunity against novel viral infections such as SARS-CoV-2. Understanding their durability and their system compartmentalization across a diverse population are critical pieces of data informing our ability to monitor seroprevalence in communities, to select plasma donors for treatment, and to design vaccines against COVID-19. We examined the stability of antibody levels over the first three months after infection in both the serum and the saliva and longitudinal sampling of serum. We observed no drastic decline in levels of anti-spike, anti-RBD or anti-NP IgG levels over a 3-month period. The same was true for the antigen-specific measurements in saliva (anti-spike and anti-RBD IgG). On the other hand, similar to other findings (*24, 25*), IgA and IgM responses to SARS-CoV-2 antigens were found to decline in both serum and saliva. In summary, our data show that a durable IgG response against SARS-CoV-2 antigens is generated in both the saliva and serum in most patients with COVID-19, and that there are some unique behaviors of the IgA response that may suggest an independent compartmentalized immune response.

Given the presence of SARS-CoV-2 RNA in saliva, it is reasonable to hypothesize that, like other viruses such as rubella (*22*), 229E alpha-coronavirus (*26*), and MERS beta-coronavirus (*27*), the mucosae and draining lymph nodes of the oro- and nasopharyngeal tracts serve as a site for initiation of an immune response to SARS-CoV-2. If so, then plasma cells (PC) that produce antibodies to SARS-CoV-2 will migrate back to the oro- and nasopharyngeal mucosae and produce antibodies that should be detectable in the saliva, a fluid that already has high levels of IgA (*28*). With time, this response will be detected in the systemic circulation, possibly due to migration of PC into new niches as we have previously described in mice (*29*). Indeed, we and two other groups have observed SARS-CoV-2 specific antibodies in saliva (*30, 31*). There are some variations between study protocols that are important to consider: Randad *et al*. applied a brush on the gum line as a means to capture IgG from the blood, heat inactivated this material, and performed Luminex to detect antigen-specific antibody levels (*31*). In contrast, our strategy was to collect saliva in a manner that best approximates the immune response that takes place in the local mucosa. In this way, our study more resembles that of Faustini *et al*, who used ELISA technology on whole saliva, amplifying the signal with an additional antibody step (*30*). Although the saliva dilutions we used closely match those of Faustini *et al*, unlike our findings, agreement between the serum and saliva for each antibody/antigen ELISA pair was less obvious in that study than in ours (*30*). Whether these discrepancies are methodological (i.e. detection of specific versus total Igs) and/or relate to the higher number of asymptomatic subjects in the Faustini *et al*. study remains to be determined.

While the sensitivity of the saliva assays was very good for anti-spike and anti-RBD IgG responses based on ROC curves, this was less true for IgA, particularly the anti-RBD IgA response. This is because some of our negative controls, irrespective of whether they were collected during the pandemic (unexposed negatives) or prior to the pandemic, exhibit AUC levels of anti-RBD IgA that approach 50% of the pooled control saliva AUC (see Figure 4H). It is unclear why this would occur for only the IgA/RBD combination. Presumably these are cross-reactive IgA that bind to SARS-CoV-2 RBD. Of interest, thus far SARS-CoV-2 neutralizing antibodies appear to have limited somatic hypermutation (*32*, *33*), suggesting that they may originate from a naïve repertoire or from B cells that have been activated in extrafollicular responses where somatic hypermutation is limited. It is tempting to speculate that these pre-existing IgA antibodies may provide some stop-gap protection against SARS-CoV-2 in the oral cavity, and if so, it is essential to ascertain their original antigenic specificity. Future work is required to confirm these results in a greater array of subjects and using different sources of RBD antigen.

Our findings that the IgG response to SARS-CoV-2 antigens is stable over a 3-month period are consistent with two other studies who likewise noted durability in the IgG response to the spike trimer (*8*, *9*). These data and ours contrast with those of Long *et al*, who showed rapid decay of antibody levels when profiling the response to a linear peptide motif of the C-terminal part of the spike protein (*10*) instead of the spike trimer used here, and it is possible that the antigen selection accounts for some of the differences. However, this does not explain discrepant results with respect to the anti-NP response in the serum, which we find also largely persisted over the 3-month period. One potential difference that could explain these divergent results is that we employed a sensitive and robust chemiluminescence-based ELISA whereas Long *et al*. employed Luminex methods.

One weakness of our study is that we have not looked beyond the day 115 PSO – our collections began in mid-March 2020 – and it is entirely plausible that antigen-specific IgG levels will eventually wane with time.

Whether this will translate to a reduction in neutralizing antibodies (nAb), as observed by Long *et al*. (*10*) but not by Wajnberg *et al*. (9), remains to be investigated in light of our own results with the snELISA. One possible explanation for the (relatively mild) decline in the nAb response observed by Long *et al*. may be due to the rapid drop in antigen-specific IgA (and potentially IgM) levels. Indeed, IgA is an important mediator of protection against gastrointestinal viruses (*34*), is essential in achieving immunity against avian viruses (*35*), and has been shown to contribute to the nAb response to SARS-CoV-2 as shown by Sterlin *et al*. (*24*). In addition, a mAb cloned from B cells derived from CoV infected humanized mice was found to provide cross-reactive neutralizing activity to SARS-CoV-2 when engineered on the IgA backbone, and this neutralizing activity was further enhanced if the IgA was co-expressed with J chain to produce dimeric IgA and secretory component to produce secretory IgA – the form of IgA that is excreted at mucosal surfaces (*36*). Although Sterlin *et al*. show that the initial IgA PB response quickly declines, IgA-producing PC have been shown to persist for decades in the gut mucosae of humans (*37*), and these will not be readily measurable in the blood. Indeed, we found that of all 3 isotypes measured, antigen-specific IgA levels in the saliva exhibited the poorest correlation with antigen-specific IgA levels in the serum. When combined with the parallel formation of re-activatable memory B cells (*38*), many of which will be tissue-resident (*21*), the host has excellent mechanisms for mounting swift and robust humoral immunity upon pathogen re-exposure that may be missed using blood-based measurements. An epidemiological study that prospectively follows confirmed COVID-19 cases for several months will determine if these immunological principals hold true in the context of SARS-CoV-2 infection.

In conclusion, our study provides evidence that the IgG response to SARS-CoV-2 spike persists in the saliva and the serum, and that this response can be correlated between the two biofluids. Given that SARS-CoV-2 initially replicates in the oro- and nasopharyngeal tracts, in the future it will be critical to characterize the nature and kinetics of salivary antibodies at the earliest time points post-infection in contact-traced individuals in order to determine if there are correlates of protection that impact viral setpoint and COVID-19 disease progression.

## Materials and Methods

*Design*. This observational study focused on monitoring the levels of antibodies to SARS-CoV-2 antigens in serum and saliva of patients with confirmed SARS-CoV-2 infection. At the onset of the study, we set to determine: 1) what are the kinetics of antibody production and decline in saliva and serum specimen from patients with COVID-19 during the first 3+ months of infection; 2) whether these levels are affected by disease severity, sex, or age; 3) whether saliva can be used as an alternative biofluid for monitoring the immune response in patients with COVID-19. Assay development was performed for each individual ELISA by assessing the classification of positives and negative samples (see definition below for serum and saliva assays) at each of the observed colorimetric or chemiluminescent values, and setting a threshold (1% for serum and 2% for saliva, respectively) for definition of positives. Irrespective of this positive/negative definition, all values are reported. The protein-based surrogate neutralization ELISA (snELISA) development and benchmarking against viral neutralization assay was described previously (*20*). Samples for profiling were recruited through the Toronto Invasive Bacterial Diseases Network in metropolitan Toronto; all samples for which a PCR positive result and for which the biofluid (serum or saliva) was available were included. Data was analyzed without exclusion of outliers to avoid biasing the study. For the saliva and snELISA assays, each sample was analyzed once, through a multipoint dilution curve; for the serum-based ELISA, a single-point ELISA was performed in duplicates, and the results averaged. No randomization was performed, since this is an observational study.

*Recruitment and participants – COVID19patients*. Acute and convalescent serum and saliva samples were obtained from patients identified by surveillance of COVID-19 (confirmed by PCR; in- and out-patients) by the Toronto Invasive Bacterial Diseases Network in metropolitan Toronto and the regional municipality of Peel in south-central Ontario, Canada (REB studies #20–044 Unity Health Network, #02-0118-U/05-0016-C, Mount Sinai Hospital). Consecutive consenting patients admitted to four TIBDN hospitals were enrolled: these patients had serum and saliva collected at hospital admission, and survivors were asked to submit repeat samples at 4–12 weeks PSO. Consecutive out-patients diagnosed at the same 4 hospitals prior to March 15^th^ and on a convenience sample of later days were approached for consent to collect serum and saliva at 4–12 weeks PSO. Patients were interviewed and patient charts reviewed to determine age, sex, symptom onset date, and disease severity (mild, moderate, and severe). For this study, disease was considered mild if it did not require hospitalization, moderate if it required hospitalization but not intensive care unit (ICU) admission, and severe if it required ICU care. Specimens were considered convalescent if they were collected less than 21 days PSO, and convalescent if they were collected 21 or more days PSO. From March 10-April 14, patients were asked to provide a 5 ml sample of saliva in a sterile specimen container, and 2.5 mls of phosphate buffered saline was added to reduce viscosity for PCR testing. From April 16^th^ on, saliva specimens were collected in Salivette® tubes (Sarstedt, Numbrecht, Germany). All specimens were aliquotted and stored frozen at −80°C prior to analysis.

Additional positive samples for test development were obtained through the Canadian Blood Services. Specimen-only serum donations were collected from individuals with a self-declared SARS-CoV-2-positive nucleic acid test. Collections occurred two weeks or more after cessation of clinical symptoms.

*Recruitment and participants – control saliva and serum*. Control saliva samples were collected from unexposed, asymptomatic individuals residing in an area of very low COVID19 case numbers (Grey County, Ontario) and throughout the Greater Toronto Area (GTA) (REB study# 23901 University of Toronto).

Control serum samples were from patients enrolled in cancer or birth cohort studies prior to COVID-19 (prior to November 2019; REB studies #01-0138-U and #01-0347-U, Mount Sinai Hospital) and archived frozen in the LTRI Biobank, or from previous studies of the immune system or systemic lupus acquired prior to November 2019 (REB studies #31593 University of Toronto, #05-0869, University Health Network).

*Study Approval*. All samples were collected after Research Ethics Board (REB) review (see Sample section above for the individual REB approval numbers). The serum ELISA assays were performed at the Lunenfeld-Tanenbaum Research Institute with Mount Sinai Hospital (MSH; Toronto, ON) Research Ethics Board (REB) approval (study number: 20-0078-E). External samples were transferred through Material Transfer Agreements as appropriate. All research has been performed in accordance with relevant guidelines and regulations. All participants have provided informed consent. The samples were de-identified prior to transfer to the assay laboratory.

*Sample collection, handling and viral inactivation – serum*. Serum (and in some cases plasma) was collected using standard procedures at the collection sites and transferred to the testing lab on dry ice. Inactivation of potential infectious viruses in plasma or serum was performed by incubation with Triton X-100 to a final concentration of 1% for 1 h prior to use (*39*).

*Antigen production – serum assays*. Spike trimer was expressed as follows: the SARS-CoV-2 spike sequence (aa 1–1208 from Genebank accession number MN908947 with the S1/S2 furin site (residues 682–685) mutated [RRAR->GGAS] and K986P / V987P stabilizing mutations was codon-optimized (*Cricetulus griseus* codon bias) and synthesized by Genscript. To stabilize the spike protein in a trimeric form, the cDNA was cloned in-frame with the human resistin cDNA (aa 23–108) containing a C-terminal FLAG-(His)_6_ tag (*Cricetulus griseus* codon bias, GenScript) into a modified cumate-inducible pTT241 expression plasmid and transfected in CHO^2353^ cells (Stuible *et al*, manuscript in preparation) followed by methionine sulfoximine selection for 14 days to generate a stable CHO pool. This CHO pool allows for cumate-inducible trimeric spike expression from the CR5 promoter as described in Poulain *et al*. (*40-42*). Cell culture was harvested 8–10 days post-cumate induction and secreted spike trimer present in the clarified medium purified by immobilized metal-affinity chromatography (Ni-Excel resin). Purified trimeric spike was buffer exchanged in PBS and store as aliquots at −80°C. The purified spike protein integrity and purity was analyzed by SDS-PAGE and analytical size-exclusion ultra-high performance liquid chromatography (SEC-UPLC). The SEC was run in PBS + 0.02% Tween-20 on an 4.6 x 300 mm Acquity BEH450 column (2.5 μm beads size; Waters Limited, Mississauga, ON) coupled to a MALS detector and the spike trimer eluted as a major (>95% integrated area) symmetrical peak of 490 kDa with less than 3% aggregates (not shown). RBD was expressed as for the saliva assay, but left non-biotinylated, as in (*20*).

Nucleocapsid^1-419^ from the pEntry-N (closed) Open Reading Frame (a kind gift from Dr. Frederick P. Roth (*43*)) was cloned into pDEST585 gift of Jim Hartley, internal ID V2097) as a HIS-GST-TEV fusion using LR-clonase. The resulting expression vector was confirmed by restriction digest, expressed in *E. coli* BL21(DE3) Codon+ cells (Agilent Technologies) and induced with 0.25 mM isopropyl 1-thio-β-D-galactopyranoside (IPTG) for 16 hours at 18°C. Harvested cells were resuspended in 20 mM HEPES pH 7.5, 400 mM NaCl, 5 mM imidazole and lysed by passage through a cell homogenizer (Avestin Inc.). Following centrifugation at 30,000g, supernatant was passed through a 0.45 μM PVDF filter and applied to a HiTrap nickel chelating HP column (GE Healthcare). Protein eluted with buffer containing 300 mM imidazole was incubated overnight with Tobacco Etch Virus (TEV) protease. Following cleavage of the His-Tag, protein was dialyzed in 20 mM HEPES pH 7.5, 50 mM NaCl and flowed over a 5 ml HiTrap nickel chelating column to remove His-GST. Nucleocapsid protein was further purified by ion exchange on a mono-S column (GE Healthcare) equilibrated in 20 mM HEPES pH 7.5, 50 mM NaCl, 1 mM DTT and eluted with a gradient to 500 mM NaCl. Purified Nucleocapsid protein was concentrated to 6 mg/mL and stored at −80°C.

*Enzyme-linked immunosorbent assays for detecting antigen-specific IgG and IgA in serum or plasma*. A manual colorimetric ELISA assay (similar to (3)) was first implemented in 96-well plates using the RBD and spike non-biotinylated antigens described here for the detection of IgG (also see (*20*)). Briefly, concentrations and incubation times were optimized to maximize the separation between anti-RBD or anti-spike trimer levels in convalescent plasma or serum from that of pre-COVID era banked serum while maintaining the required levels of antigens as low as possible. 75 ng and 200 ng of RBD and spike, respectively, were first adsorbed onto 96-well clear Immulon 4 HBX (Thermo Scientific, #3855) plates in PBS overnight at 4°C, then washed three times with 200 μl PBS+ 0.1% Tween-20 (PBS-T; Sigma). Plates were blocked with 3% w/v milk powder (BioShop Canada Inc., #ALB005.250, lot #9H61718) in PBS for 1–2 hr and washed three times with 200 μl PBS-T. Patient samples (pre-treated with 1% final Triton X-100 for viral inactivation) diluted 1:50 in PBS-T containing 1% w/v milk powder were then added to the plates and incubated for 2 hr at room temperature (50 μl total volume): technical duplicates were performed unless otherwise indicated. Positive and negative control recombinant antibodies and serum samples were added to each plate to enable cross-plate comparisons. Wells were washed three times with 200 μl PBS-T. Goat anti-human anti-IgG (Goat anti-human IgG Fcy −HRP, Jackson ImmunoResearch, #109-035-098) at a 1:60,000 dilution (0.67 ng/well) in 1% w/v milk powder in PBS-T was added and incubated for 1 hr. Wells were washed three times with 200 μl PBS-T, and 50 μl of 1-Step™Ultra TMB-ELISA Substrate Solution (ThermoFisher, #34029) was added for 15 min at room temperature and the reaction was quenched with 50 μl stop solution containing 0.16N sulfuric acid (ThermoFisher, #N600). The plates were read in a spectrophotometer (BioTek Instruments Inc., Cytation 3) at 450 nm. For all ELISA-based assays, raw OD values had blank values subtracted prior to analysis. All data were normalized to the positive serum control (single point) on each plate and expressed as a ratio to this control. The assay performance was assessed by precision-recall analysis of ratio-expressed values (Table 1; Supplemental Figure 1, 3).

The assay was then re-designed to be conducted in a customized robotic platform using a 384-well plate format, first by simply scaling down the volume/amounts used, and then switching to a chemiluminescent substrate for detection, and re-optimizing the amounts per well of antigens and secondary antibodies’ dilutions to use. A chemiluminescent substrate is ideally-suited for automated ELISAs, because it offers a higher sensitivity and a better dynamic range than standard colorimetric assays. Furthermore, the reaction does not need to be stopped (e.g. with robotics-incompatible acids) and the luminescence signal is stable for at least 60 min. For all steps, liquid dispensers (Beckman Biomek NXp or ThermoFisher Multidrop Combi) and washer (Biotek 405 TS/LS LHC2) were used on a F7 robotic platform available at the Network Biology Collaborative Centre (nbcc.lunenfeld.ca). Each step of the methods to evaluate the different antigen and antibody class combinations were optimized and routine quality control tests were performed for all dispensing steps.

For automated ELISAs, LUMITRAC 600 high-binding white polystyrene 384-well microplates (Greiner Bio-One, through VWR #82051-268) were pre-coated overnight with 10 μl/well of RBD (25 ng) or spike (50 ng). Next day, the wells were washed 4 times (a BioTek washer is used for all washing steps, and all washes are performed with 100 μl PBST). Wells were blocked for 1 hr at room temperature in 80 μl 5% Blocker™ BLOTTO (Thermo Scientific, #37530), then washed 4 times. 10 μl Triton X-100 inactivated serum (or plasma) samples diluted 1:40 in 1% BLOTTO in PBS-T were added to each well from 96-well sample source plates and incubated for 2 hr at room temperature. Positive and negative controls used on each plate are described below. After washing 4 times, 10 μl of one of the following secondary antibodies (all from Jackson ImmunoResearch) diluted in 1% BLOTTO in PBS-T were added at the indicated concentrations followed by incubation for 2 hr at room temperature: Goat anti-human IgG Fcy – HRP (#109-035-098; 1:40,000 or 0.2 ng per well), Goat anti-human IgM Fc5u – HRP (#109035-129; 1:12,000 or 0.66 ng per well) or Goat anti-human IgA a chain – HRP (#109-035-127; 1:10,000 or 0.8 ng per well). After 4 washes, 10 μl of SuperSignal ELISA pico Chemiluminescent substrate (diluted 1:4 in water) was added, followed by a short mix for 10s at 900 rpm, and incubation at room temperature for 5 min. Luminescence was read on an EnVision (Perkin Elmer) plate reader at 100 ms/well using an ultra-sensitive luminescence detector. All automated assays were performed in biological duplicates, processed on different days. Blank values were subtracted for all raw reads prior to data analysis, and the values were expressed as a ratio of the positive reference serum pool on the same plate (see below).

Quality controls and normalization of the samples in the automated assays were as follows: a standard curve with recombinant antibodies reacting to spike RBD or spike S1 was included on each plate. Antibodies used for the standard curves were: Human anti-spike S1 IgG (A02038, GenScript), anti-spike S1 IgM (A02046, GenScript) and Ab01680 anti-spike IgA (Ab01680-16, Absolute Antibody), all used at 0.5 to 10ng per well. Negative antibody controls were immunoglobulins from human serum (I4506 human IgG, I8260 human IgM, and I4036 human IgA, from Millipore-Sigma). A positive and negative control pool of 4 patient samples each was created and added in each plate at a single point concentration for normalization. For all assays, a standard curve is generated by first plotting the mean of the blank-subtracted recombinant antibodies, plotted against antibody amounts (in ng), and the linearity of the curve and comparison to previous runs is assessed, alongside the confirmation that the positive and negative pool sample fall within the expected range of the standard curve [%CV should be 10–15% or less].

The surrogate neutralization ELISA (snELISA) was performed manually as described recently (*20*).

*Surrogate Enzyme-linked immunosorbent assays for detecting antigen-specific IgG, IgA and IgM in serum or plasma*.

*Sample collection and handling – saliva study*. With the exception of some samples that were acquired early on in the pandemic (cohort 1), Salivette® tubes were used to collect samples according to manufacturer instructions (Sarstedt, Montreal, Quebec). These tubes include a cotton swab that participants are instructed to chew for set amount of time. The swab is then transferred into an inner tube which is then inserted into an outer tube that catches liquid saliva upon centrifugation at 1000 x g for 3 minutes. Salivary flow was controlled by establishing a fixed amount of collection time (2 minutes) for each subject as previously recommended (*20, 44*). For the early pandemic subjects that were not given salivettes and used in our pilot study (cohort 1), these subjects expectorated directly into a 15 mL conical tube containing 2.5 mL of phosphate-buffered saline (PBS). Prior to saliva collection, healthy subjects confirmed they had fasted, refrained from taking oral medication, and had not brushed their teeth for a minimum of 30 minutes.

*Viral inactivation in saliva samples*. Following centrifugation, all saliva samples, regardless of their SARS-CoV-2 PCR status, underwent viral inactivation by treating with Triton® X-100 (BioShop, CAT# TRX506.100). 10% Triton X-100 (diluted 1:10 from stock) was added to all samples to a final dilution of 1% Triton X-100 and incubated for 1 hour at room temperature. Inactivated samples were immediately frozen and stored at −80° C. Heat inactivation for 30 minutes at 65°C was found to destroy the IgG and IgA signal against RBD and was therefore not used (Supplemental Figure 5). The efficiency of virus inactivation in a saliva medium is shown in Table S3. Specifically, we assessed the treatment of saliva collected from healthy individuals using two different methods (Salivette vs. direct saliva collection into a tube). These samples were spiked with known amounts of SARS-CoV-2 viral stock and then treated with 1% Triton X-100 for 30 minutes, 1 hour or 2 hours. Vero-E6 cells (ATCC® CRL-1586™) were used to determine outgrowth of virus. Cells were maintained in Dulbecco’s Modified Eagle’s Media (DMEM) supplemented with L-glutamine, penicillin/streptomycin and 10% fetal bovine serum (FBS). SARS-CoV-2 virus (isolate SB3) was isolated in-house (*45*). Briefly, viral stocks were created after isolation of virus from a clinical sample in Toronto, Ontario, Canada. Viral stock was expanded using Vero E6 as previously described such that stored aliquots of stock contain 2% FBS. Initial experiments were done with Triton X-100 (Sigma-Aldrich) serially diluted and applied to Vero-E6 cells in 96-well flat bottom plates to determine the minimum concentration required to prevent toxicity to cells. Furthermore, we have also determined if neat saliva itself could be cytotoxic to Vero-E6 cells by providing healthy donor saliva alone or treated with Triton X-100 ranging from final Triton-X100 concentration of 0.03%, 0.01%, 0.001% and 0.0001% (v/v). Since initial Triton X-100 experiments showed that toxicity is averted at 0.03% (v/v), we proceeded to use this concentration as the point of dilution to prevent any Triton X-100 mediated toxicity.

*Antigen production – saliva assay*. The expression, purification and biotinylation of the SARS-CoV-2 RBD and spike ectodomain were performed as recently described (*20, 44*). The human codon optimized cDNA of the SARS-CoV-2 spike protein was purchased from Genscript (MC_0101081). The soluble RBD (residues 328–528, RFPN…CGPK) was expressed as a fusion protein containing a C-terminal 6xHis tag followed by an AviTag. The soluble trimeric spike protein ectodomain (residues 1–1211, MFVF…QYIK) was expressed with a C-terminal phage foldon trimerization motif followed by a 6xHis tag and an AviTag. To help stabilize the spike trimer in its prefusion conformation, residues 682–685 (RRAR) were mutated to SSAS to remove the furin cleavage site and residues 986 and 987 (KV) were each mutated to a proline residue (*46*). Stably transfected FreeStyle 293-F cells secreting the RBD and soluble spike trimer were generated using a previously reported piggyBac transposon-based mammalian cell expression system (*47*). Protein production was scaled up in 1L shake flasks containing 300 mL FreeStyle 293 medium. At a cell density of 10^6^ cells/mL, 1 μg/mL doxycyline and 1 μg/mL Aprotinin were added. Every other day 150 mL of medium was removed and replaced by fresh medium. The collected medium was centrifuged at 10000 x g to remove the cells and debris and the His-tagged proteins were purified by Ni-NTA chromatography. The eluted protein was stored in PBS containing 300 mM imidazole, 0.1% (v/v) protease inhibitor cocktail (Sigma, P-8849) and 40% glycerol at −12 °C. Shortly before use, the RBD and spike proteins were further purified by size-exclusion chromatography on a Superdex 200 Increase (GE healthcare) or Superose 6 Increase (GE healthcare) column, respectively. Purity was confirmed by SDS-PAGE (not shown). For the spike protein, negative stain electron microscopy was used show evidence of high-quality trimers (not shown). The Avi-tagged proteins, at a concentration of 100 μM or less, were biotinylated in reaction mixtures containing 200 μM biotin, 500 μM ATP, 500 μM MgCl_2_, 30 μg/mL BirA, 0.1% (v/v) protease inhibitor cocktail. The mixture was incubated at 30 °C for 2 hours followed by size-exclusion chromatography to remove unreacted biotin.

*Enzyme-linked immunosorbent assays for detecting total IgA, IgG and IgM in saliva*. Quantitative total IgA, IgG, and IgM analyses were performed on the same samples used for detection of anti-RBD and anti-spike Ig described below. Anti-human Ig antibody (Southern Biotech, 2010–01) diluted 1:1000 in PBS was added to 96-well Nunc MaxiSorp™ plates (ThermoFisher, 44-2404-21). PBS alone was added to control wells. Plates were allowed to coat overnight at 4°C. Following coating, plates were blocked using 5% BLOTTO for 2 hours at 37°C. Samples were diluted in 2.5% BLOTTO. Standards (purified IgA, IgG and IgM purchased from Sigma-Millipore (IgA, I4036, IgG, I2511, and IgM, I8260) were prepared in 2.5% BLOTTO ranging from 100 ng/mL 3.125 ng/mL. Upon discarding the blocking solution from the plate, samples and standards were immediately transferred to wells and incubated for 2 hours at 37°C. Following incubation, wells were washed with 200 μl of PBS-T. HRP-conjugated secondary antibodies against IgA, IgG, and IgM (goat anti-human IgA- and IgG-HRP, Southern Biotech, IgA: 2053–05, IgG: 2044–05, IgM: 2023–05) were added to the appropriate wells at 1:1000 in 2.5% BLOTTO and incubated for 1 hour at 37°C. Development of the plate was done by adding 50 μL of 3,3’,5,5’-tetramethylbenzidine (TMB) Substrate Solution (ThermoFisher, 004021–56) onto plates. Reaction vas then stopped by adding 50ul/well of 1N H_2_SO_4_ Optical density (OD) was read at a wavelength of 450 nm on a spectrophotometer (OD_450_). A four-parameter logistic curve was used to determine the line of best fit for the standard curve, and sample Ig quantities were interpolated accordingly to determine final concentrations in μg/ml. The few samples from patient or control groups that exhibited quality control issues (extremely low to negative IgA levels) were excluded from further analysis.

*Enzyme-linked immunosorbent assay for detecting albumin in saliva*. Salivary albumin was measured for Cohort 1 using a purchased Human Albumin ELISA Kit (Abcam, ab108788). Assay was performed according to manufacturer instructions included with the kit.

*Enzyme-linked immunosorbent assays for detecting antigen-specific IgG, IgA and IgM in saliva*. 96-well plates pre-coated with streptavidin (ThermoFisher, 436014) were used for all assays. Without the biotin-streptavidin system, the anti-S/RBD IgG, IgA, and IgM signals obtained from COVID-19 patient saliva were undetectable. Based on titrations of antigens using saliva from convalescent COVID-19 patients, 100ng of biotinylated-RBD and 1μg of biotinylated S proteins were applied to the appropriate wells one day prior to starting the assay (see Supplemental Figure 5 for RBD titration, spike titration not shown). Control wells of sterile PBS rather than biotinylated antigen were reserved for each patient and control sample. A few wells with the biotinylated antigen but with no sample added were reserved as negative internal controls for the reagents on the assay. Plates were incubated overnight at 4°C to allow sufficient coating of the antigen. 200 μL of 5% BLOTTO (5% w/vol skim milk powder (BioShop, CAT# SKI400.500) in sterile PBS) was subsequently added to each well to prevent non-specific interactions, followed by a 2-hour incubation at 37°C. Blocking solution was discarded immediately from plates prior to addition of samples to wells. Newly thawed saliva samples were centrifuged at 8000 rpm for 4 minutes, and appropriately diluted using 2.5% BLOTTO. To reduce anti-streptavidin reactivity in the saliva, diluted samples were applied to streptavidin-coated plates with no antigen and allowed to incubate for 30 minutes at 37°C. Subsequently 50 μL of samples were transferred from the pre-adsorption plate into antigen-coated plates and incubated for 2 hours at 37°C. PBS+0.05% Tween 20 (BioShop, CAT# TWN510) (PBS-T) was used for washing plates between steps. Horseradish peroxidase (HRP)-conjugated Goat anti human-IgG, IgA, and anti-IgM secondary antibodies (Southern Biotech, IgG: 2044–05, IgA: 2053–05, IgM: 2023–05) were added to wells at dilutions of 1:1000, 1:2000 and 1:1000 in 2.5% BLOTTO, respectively, and incubated for 1 hour at 37°C. Development of the plates was performed as described in the section above. For cohort 1, because some samples had been collected in cups and were therefore diluted, normalization to a separate variable was performed. The resulting OD from antigen-specific IgA and IgG was subtracted from the OD for the PBS control wells for each sample and subsequently normalized to albumin levels or total IgA and IgG levels, respectively (see below). IgM was not calculated for cohort 1 due to lack of remaining sample from the COVID-19 patients. For cohort 1, raw OD_450_ measurements obtained from PBS-coated wells corresponding to each sample diluted at 1/5 (“background signal”) was subtracted from readings obtained from antigen-coated wells at each of three dilutions (1/5, 1/10). Data was normalized to the total IgG, total IgA, or albumin content in each saliva sample. A small number of samples (n=9 from negative controls and n=4 from patients) exhibited high OD values that did not titrate and coincided with high OD levels when plated without antigen (PBS control). These were excluded from the analysis. For cohort 2, raw OD_450_ measurements obtained from PBS-coated wells corresponding to each sample diluted at 1/5 (“background signal”) was subtracted from readings obtained from antigen-coated wells at each of three dilutions (1/5, 1/10, 1/20). For each plate, a sample of pooled saliva from COVID-19 acute and convalescent patients was likewise plated at 1/5 with no antigen (PBS control), as well as with antigens at 1/5, 1/10 and 1/20. The area under the curve (AUC) was calculated based on the background subtracted values from all three dilutions for each sample, was normalized to the AUC for the pooled positive control saliva and expressed as a percentage (Supplemental Figure 6A). By using the same positive control that we ran in every single plate, we determined that intra-assay precision was always greater than 90% between wells. Reproducibility between plates was determined by a coefficient of variation of less than 10% through all the plates. A small number of samples (n=6 from negative controls and n=2 from patients) exhibited high OD values that did not titrate and coincided with high OD levels when plated without antigen (PBS control) (Supplemental Figure 6B). These were excluded from the final analysis.

*Receiver-Operating Characteristic* (*ROC*) *curves*. For serum and plasma sample analysis, samples acquired prior to November 2019 (pre-COVID) were labeled true negatives while convalescent samples from patients with PCR-confirmed COVID19 were labelled true positives. For saliva samples, all samples from patients with PCR-confirmed COVID19 collected more than 10 days PSO were considered true positives, and saliva collected before 2020 and from unexposed, asymptomatic individuals in March of 2020 were labeled true negative for ROC analysis. Ratio-converted ELISA reads (colorimetric or chemiluminescent) were used for ROC analysis in the easyROC webtool (v 1.3.1) with default parameters (https://journal.r-project.org/archive/2016/RJ-2016-042/index.html). Non-parametric curve fitting was applied alongside DeLong’s method for standard error estimation and confidence interval generation.

### Statistical analysis

For both antigen-specific and total IgA, IgG and IgM readouts in saliva, raw OD_450_ measurements obtained from PBS-coated wells corresponding to each sample (“background signal”) was subtracted from readings obtained from antigen-coated or anti-human Ig-coated wells. Total IgA, IgG and IgM quantification were determined relative to standard wells present on each plate. A four-parameter logistic curve was used to determine the line of best fit for the standard curve, and sample Ig quantities were interpolated accordingly, using Prism (GraphPad), Version 8.3. The resulting OD from antigen-specific IgA and IgG were subsequently normalized to total IgA and IgG, respectively, for cohort 1. For cohort 2, the resulting antigen-specific IgA, IgG and IgM OD values across three dilutions were used to calculate the AUC for each sample. The sample AUC was normalized to the AUC of a positive pool of saliva samples used as an internal standard across all plates and the values were expressed as a percentage. For serum, raw OD_450_ measurements for IgG, IgA and IgM on spike, RBD and NP from either the manual or automated platforms were subtracted from wells coated with PBS. A pool of serum samples that previously exhibited high levels of IgGs to all antigens was used as an internal standard across all plates, and a relative ratio between blank-adjusted OD_450_ measurements of patient samples to the OD_450_ measurements of this positive pooled standard are reported. Serum data were analyzed in R using version 4.0.1. Median antibody levels between negative and positive subject groups (saliva) or negative, acute and convalescent subject groups (blood) were compared using Mann Whitney U tests. These analyses were performed in Prism (GraphPad), Version 8.3.

The relationship between time PSO and antibody levels in the convalescent period was examined in multivariable linear regression models that adjusted for age, sex, and disease severity. For serum samples, seven multivariable linear regression models were constructed (one for each of anti-RBD IgA, anti-S IgA, anti-RBD IgG, anti-S IgG, anti-RBD IgM, anti-spike IgM, neutralizing antibody). Generalized estimating equations were used (proc genmod in SAS with exchangeable correlation matrices) to account for patient-level clustering. Antibody levels were transformed as appropriate to achieve heteroscedasticity, and the variance inflation factors for all covariates confirmed to be <5 to verify absence of multicollinearity. For saliva samples, six multivariable linear regression models were similarly constructed; however, only the first convalescent sample for each patient was included in the analysis (proc glm in SAS).

## Data Availability

All the aggregate data is available in the manuscript; access to the raw, unprocessed data is available upon request.

## Supplementary Materials

Fig. S1. Development and validation of manual colorimetric and automated chemiluminescent assays for monitoring RBD and spike trimers antibodies in serum or plasma.

Fig. S2. Correlations between antibody levels and day of symptom onset to sample collection.

Fig. S3. IgG and IgA responses to the Nucleocapsid antigen of SARS-CoV2 in serum.

Fig. S4. Effect of heat versus detergent inactivation of saliva samples on the detection of anti-RBD antibodies in a manual, colourimetric ELISA.

Fig. S5. IgG and IgA levels against SARS-CoV-2 antigens in the saliva of cohort 1.

Fig. S6. Strategy for %AUC calculations in antigen-specific saliva assay.

Fig. S7. Validation of manual colourimetric assays for monitoring RBD and spike trimers antibodies in saliva.

Table S1. ROC statistics table for ELISAs conducted on blood-derived samples.

Table S2. Testing effect of Triton-X treatment on saliva samples.

Table S3. ROC statistics table for ELISAs conducted on saliva samples.

**References and Notes:**

## Acknowledgments

We thank Janet McManus at Canadian Blood Services for her technical and logistical expertise, Dzana Dervovic, Cassandra Wong, Mark Jen and Elizabeth Rubie for help with serum-based ELISAs, Joan Wither for the lupus patient samples, Frederick P. Roth for the *SARS-CoV-2* Nucleocapsid entry vector and Jim Hartley for bacteria expression vectors. We thank research associates of the Toronto Invasive Bacterial Diseases Network and Antonio Estacio, Megan Buchholz, Patti Cheatley, Rebecca Zavalunov and Katerina Pavenski for specimen collection. We thank Jeff Browning (Boston University) for critical input into the saliva experiments.

## Supplementary Materials

**Supplemental Figure 1.**
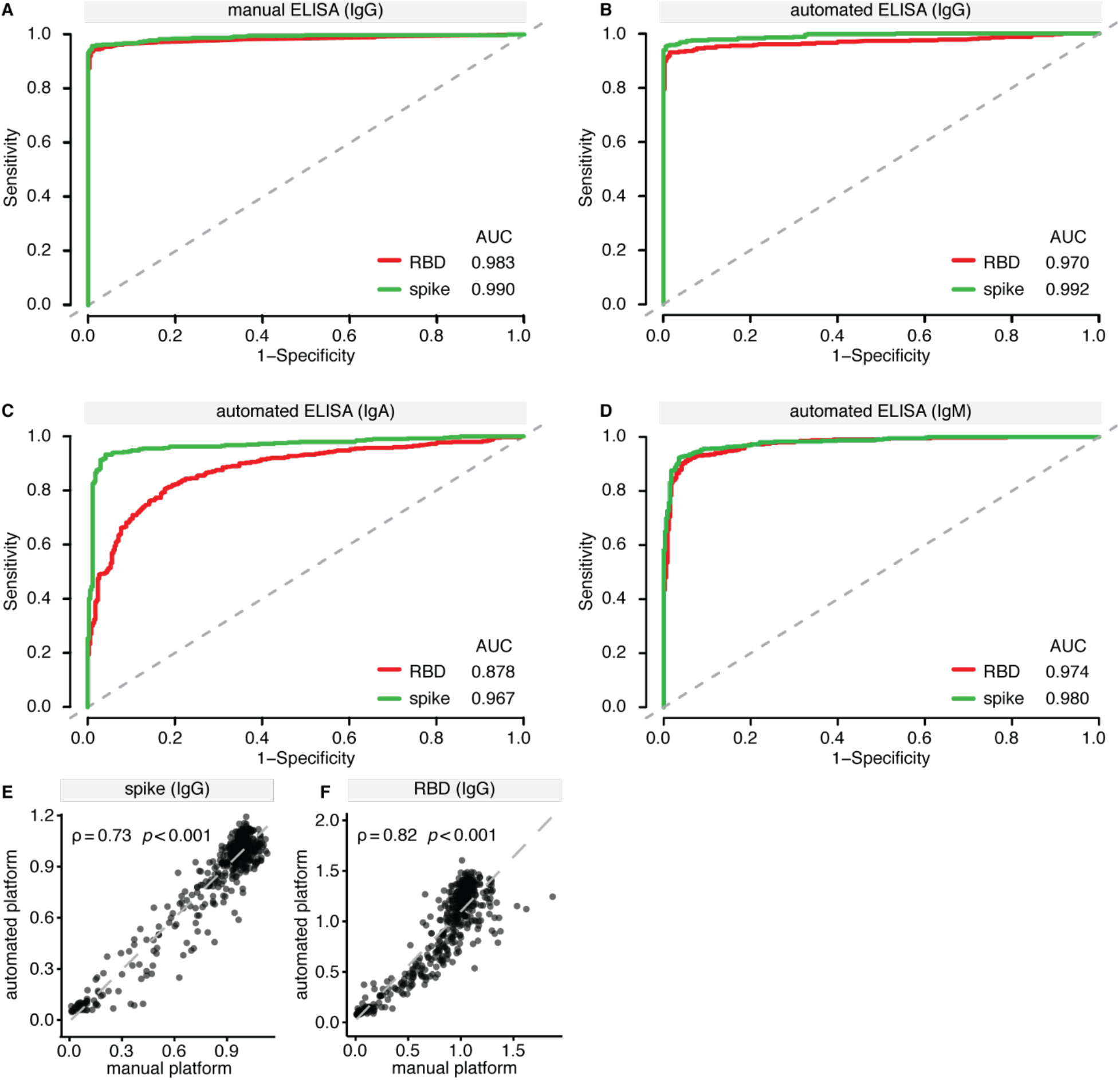
Development and validation of manual colorimetric and automated chemiluminescent assays for monitoring RBD and spike trimers antibodies in serum or plasma. (**A-D**) Receiver operating characteristic (ROC) analysis of serum/plasma assays. ROC curves generated for ELISAs conducted on serum (manual IgG and automated IgG/IgA/IgM platforms) on spike and RBD. Samples used for the profiling are listed in Supplemental Table 1. (**E-F**) Spearman correlation between IgG readouts of manual (colorimetric, 96 wells) and automated (chemiluminescent, 384 wells) assays using spike and RBD.

**Supplemental Figure 2.**
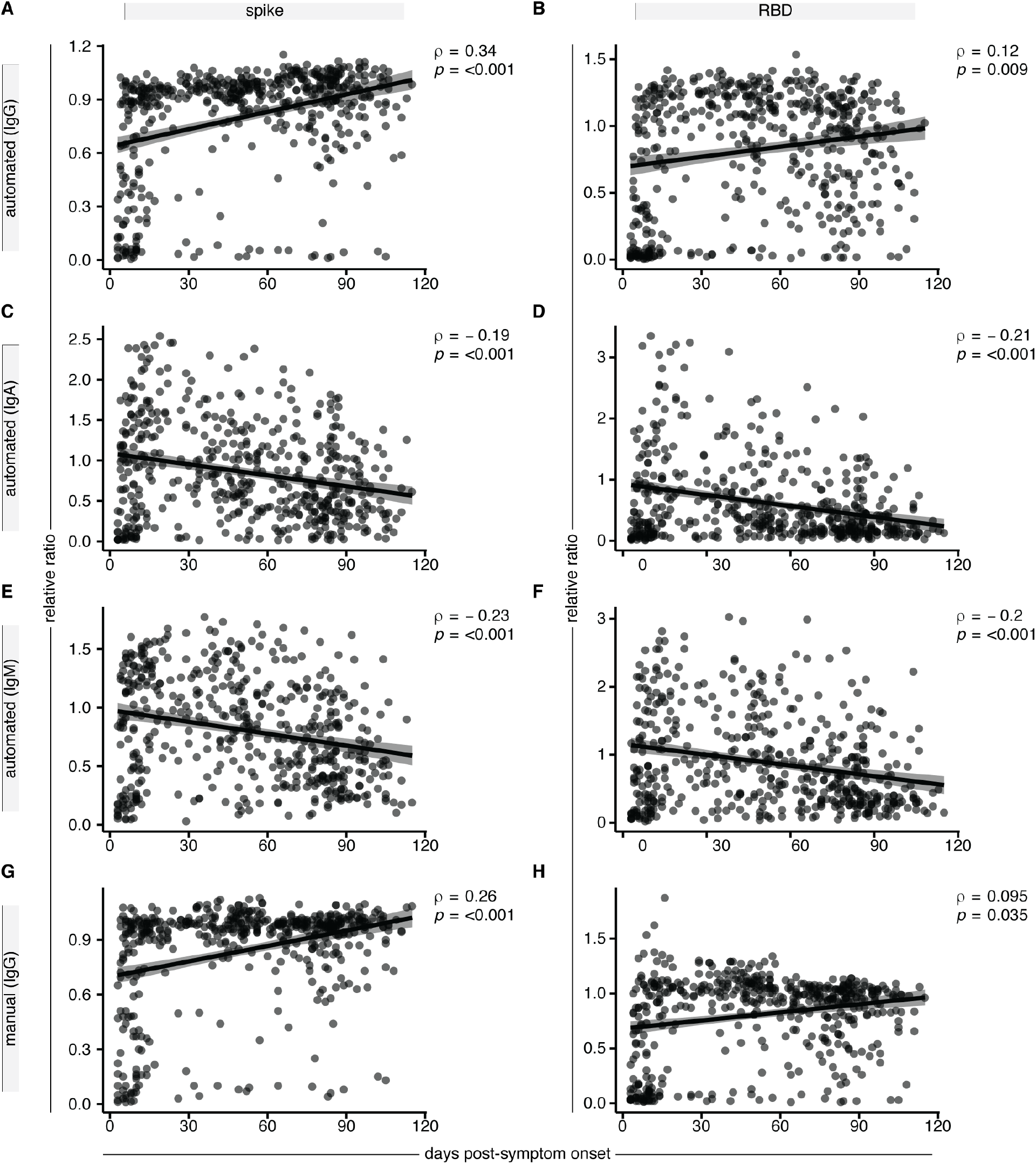
Correlations between antibody levels and day of symptom onset to sample collection. (**A-D**) Spearman rank correlations between ELISA results from the automated chemiluminescent platform for the indicated antigens (spike trimer left column; RBD right column), and immunoglobulins (**A-B**; IgG; **C-D**; IgA). (**E-F**) Validation of the trends observed in the automated platform on the manual colorimetric platform for the IgG response to spike and RBD. The shaded area represents the 95% confidence interval.

**Supplemental Figure 3.**
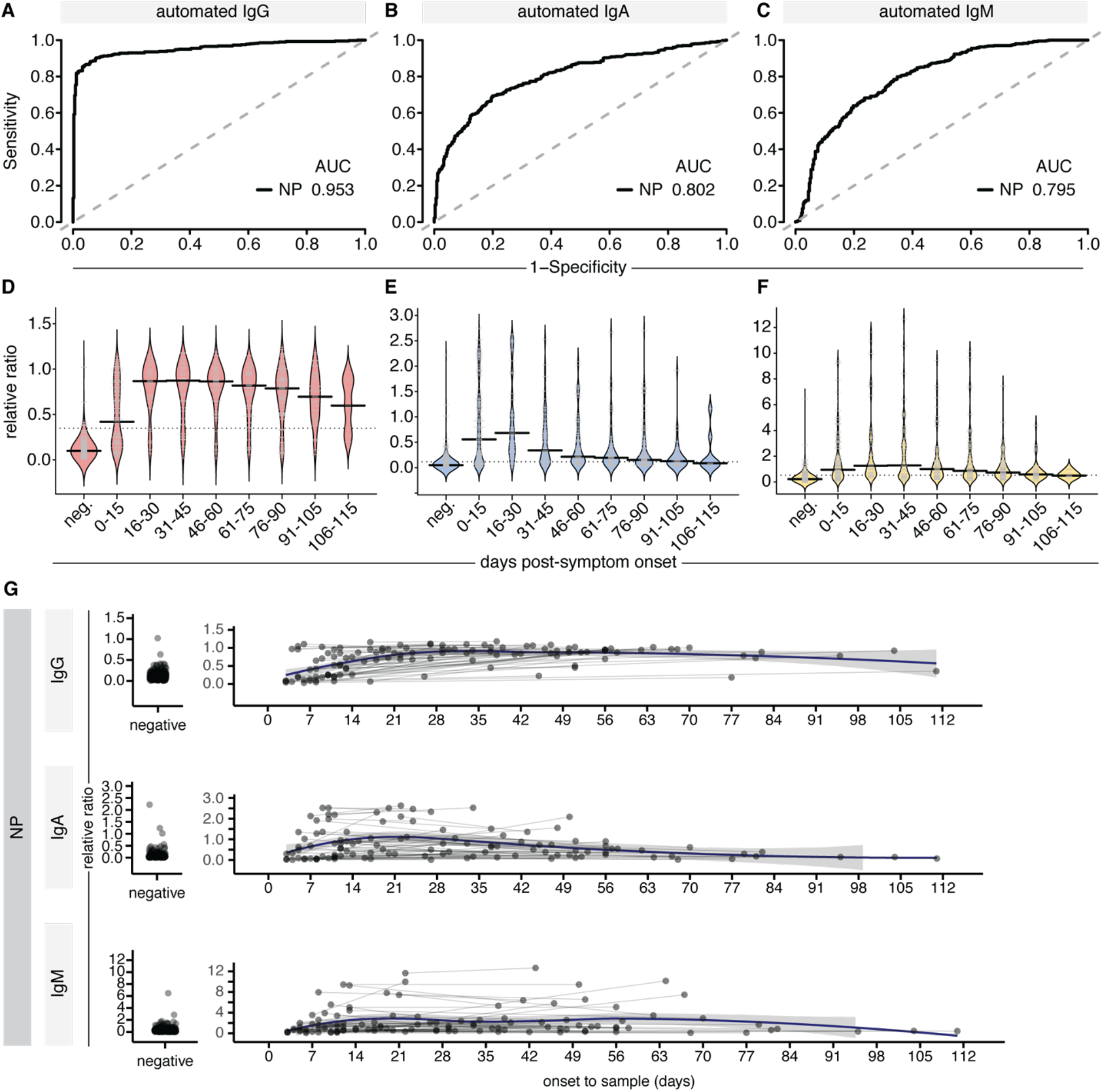
IgG and IgA responses to the Nucleocapsid antigen of SARS-CoV2 in serum. (**A-C**) Receiver operating characteristic (ROC) analysis of serum/plasma assays for NP (as in Supplemental Figure 1). (**D-F**) Binned relative ratios of IgG, IgA and IgM to NP, displayed as bean plots (see Figure 2). Solid bars denote the median and dotted line represents the median across all samples used in the plot. (**G**) Longitudinal profiling of the antibody response to NP in patients profiled twice, with non-parametric loess function shown as the blue line, with the grey shade representing the 95% confidence interval (also see Figure 3).

**Supplemental Figure 4.**
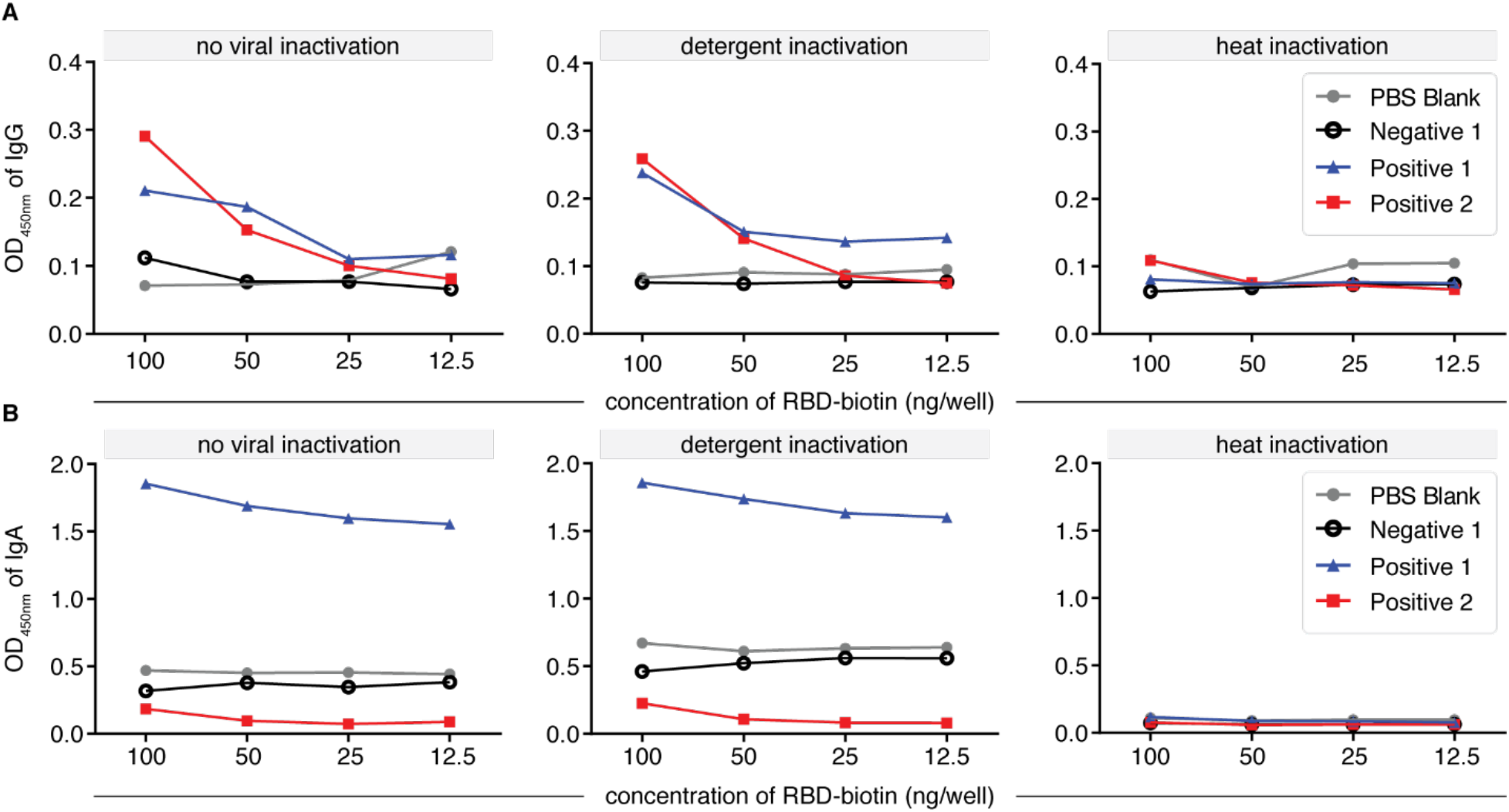
Effect of heat versus detergent inactivation of saliva samples on the detection of anti-RBD antibodies in a manual, colourimetric ELISA. (**A**) Anti-RBD IgG levels expressed as raw OD values in heat versus detergent inactivated samples across a titration of RBD-biotin levels. (**B**) Anti-RBD IgA levels expressed as raw OD values in heat versus detergent inactivated samples across a titration of RBD-biotin levels.

**Supplemental Figure 5.**
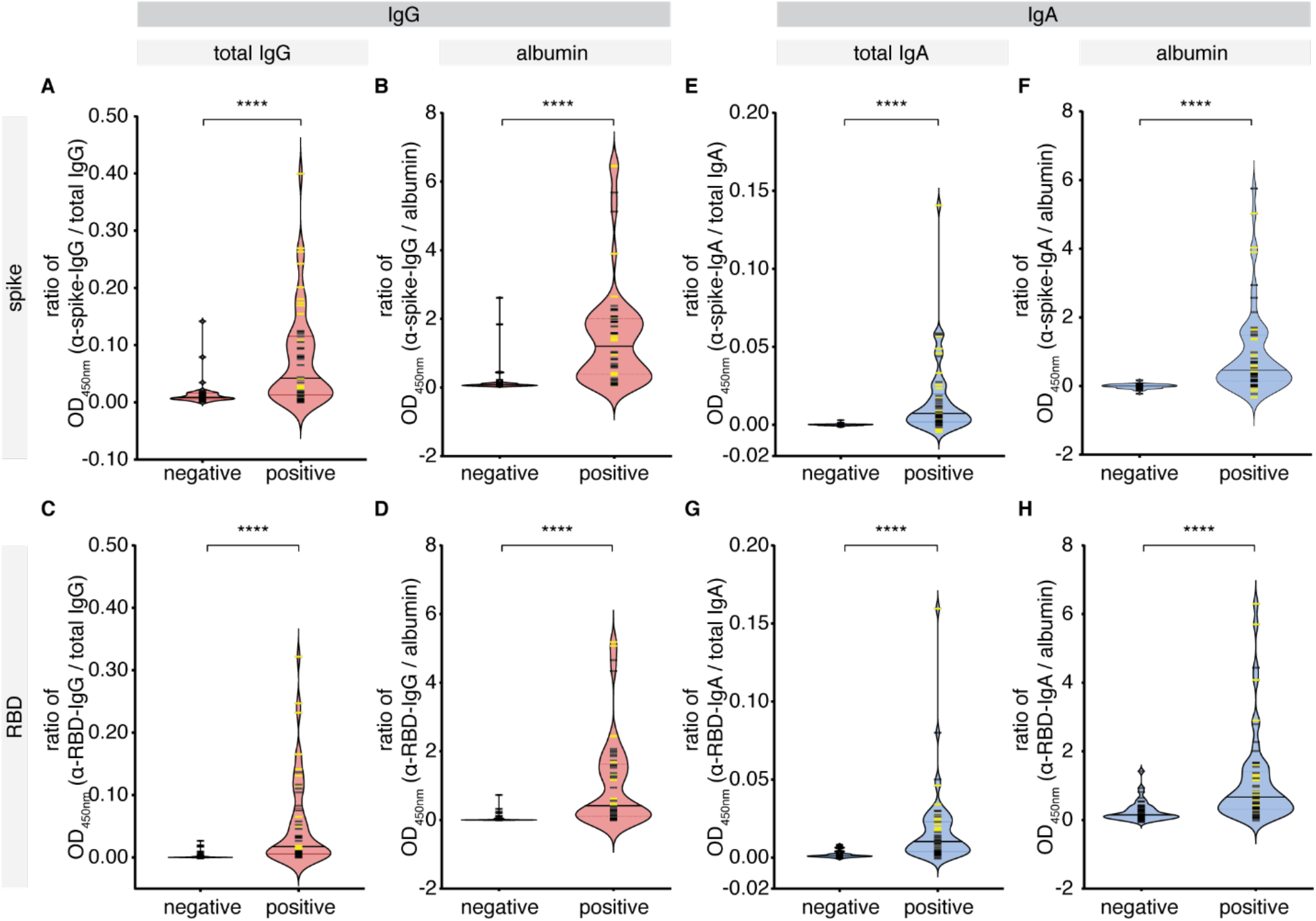
IgG and IgA levels against SARS-CoV-2 antigens in the saliva of cohort 1. A pilot cohort of COVID-19 patients was tested for the presence of IgG and IgA antibodies to SARS-CoV-2 spike and RBD antigens in the saliva, comparing with age- and sex-matched unexposed negative controls collected locally. (**A-D**) Antigen-specific (anti-spike, anti-RBD) IgG levels normalized by total IgG or albumin. (**E-H**) Antigen-specific (anti-spike, anti-RBD) IgA levels normalized by total IgA or albumin. Yellow bars denote saliva samples collected at an unknown dilution. Solid bars denote the median. Mann-Whitney U test for significance was performed. **** = p < 0.0001.

**Supplemental Figure 6.**
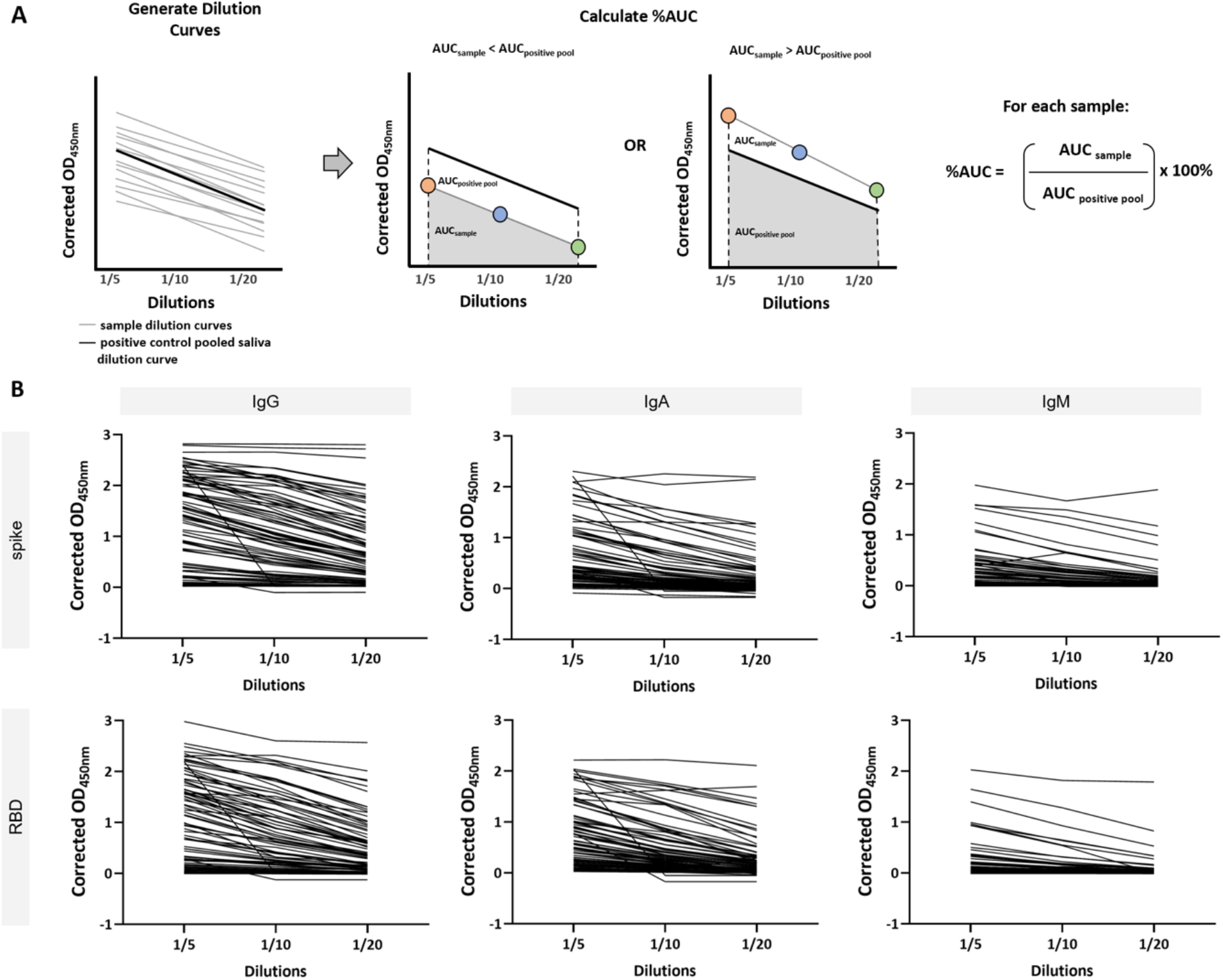
Strategy for %AUC calculations in antigen-specific saliva assay. (**A**) Schematic representation of %AUC calculation. For every sample, three dilutions are run at 1/5, 1/10 and 1/20 and the raw OD_450nm_ is determined. These OD values are corrected by background subtraction and the AUC is calculated. The AUC of each sample is normalized to the AUC of a pool positive saliva control and the percentage is expressed. (**B**) Background corrected OD_450nm_ dilution curves for COVID-19 positive patient saliva samples are expressed for all 6 antigen-antibody readouts.

**Supplemental Figure 7.**
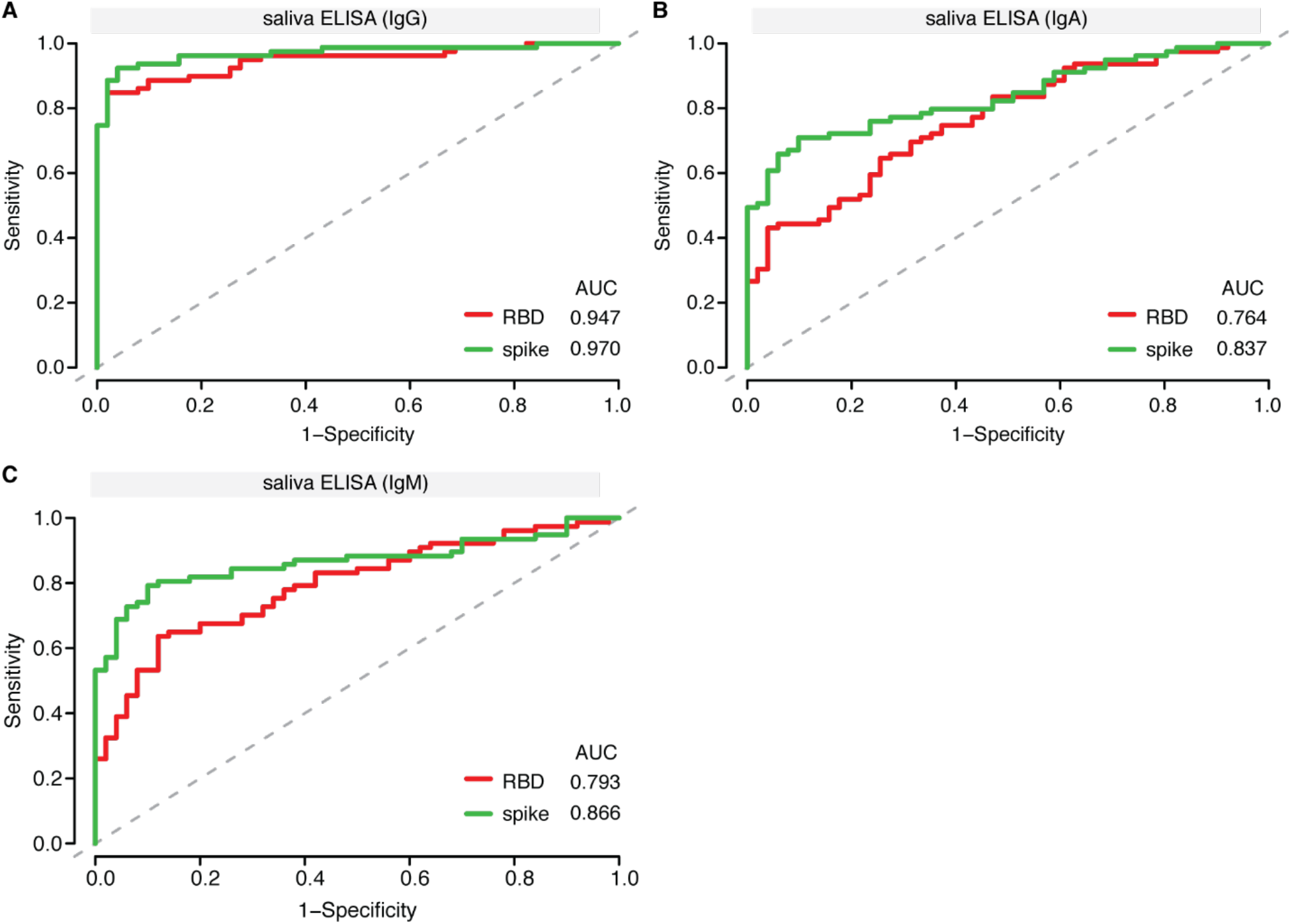
Validation of manual colourimetric assays for monitoring RBD and spike trimers antibodies in saliva. (**A-C**) Receiver operating characteristic (ROC) analysis of ELISA assays conducted on saliva (manually conducted IgG/IgA/IgM) on spike and RBD. Samples used for the profiling are listed in Supplemental Table 3.

**Table S1.** ROC statistics table for ELISAs conducted on blood-derived samples*.

|  | *Manual IgG* |  |  | *Automated IgG* |  |  | *Automated IgA* |  |  | *Automated IgM* |  |  |
| --- | --- | --- | --- | --- | --- | --- | --- | --- | --- | --- | --- | --- |
| Marker | Spike | RBD | NP | Spike | RBD | NP | Spike | RBD | NP | Spike | RBD | NP |
| AUC | 0.990 | 0.983 | 0.975 | 0.992 | 0.970 | 0.953 | 0.967 | 0.878 | 0.802 | 0.980 | 0.974 | 0.795 |
| SE.AUC | 0.0033 | 0.0047 | 0.0056 | 0.0025 | 0.0068 | 0.0079 | 0.0068 | 0.0127 | 0.0160 | 0.0043 | 0.0051 | 0.0166 |
| Lower Limit | 0.984 | 0.974 | 0.964 | 0.987 | 0.957 | 0.937 | 0.954 | 0.853 | 0.771 | 0.971 | 0.964 | 0.762 |
| Upper Limit | 0.997 | 0.993 | 0.986 | 0.997 | 0.984 | 0.968 | 0.981 | 0.903 | 0.834 | 0.988 | 0.984 | 0.827 |
| z | 148.22 | 102.12 | 85.28 | 199.89 | 69.10 | 57.20 | 69.01 | 29.78 | 18.86 | 110.61 | 93.12 | 17.81 |
| p-value | 0 | 0 | 0 | 0 | 0 | 0 | 0 | 0 | 0 | 0 | 0 | 0 |
| *ROC Coordinate points at <1% FDR* |  |  |  |  |  |  |  |  |  |  |  |  |
| Marker | Spike | RBD | NP | Spike | RBD | NP | Spike | RBD | NP | Spike | RBD | NP |
| Cutpoint | 0.35 | 0.19 | 0.49 | 0.124 | 0.244 | 0.53 | 0.735 | 0.616 | 0.559 | 0.437 | 0.382 | 2.79 |
| FPR | 0.0088 | 0.0088 | 0.0088 | 0.0088 | 0.0088 | 0.0088 | 0.0088 | 0.0088 | 0.0088 | 0.0088 | 0.0088 | 0.0088 |
| TPR | 0.956 | 0.938 | 0.828 | 0.955 | 0.913 | 0.754 | 0.445 | 0.301 | 0.223 | 0.726 | 0.657 | 0.075 |
| *Samples used to generate ROC curves* |  |  |  |  |  |  |  |  |  |  |  |  |
| Sample Cohort | No. Samples |  |  |  |  |  |  |  |  |  |  |  |
| COVID-19 Patient Samples | 402 |  |  |  |  |  |  |  |  |  |  |  |
| Pre-COVID-19 Negative Controls | 339 |  |  |  |  |  |  |  |  |  |  |  |

\* Sensitivity values reported in the manuscript are based on the coordinate points at a false positive rate of <1%.

**Table S2.** Testing effect of Triton-X treatment on saliva samples*.

|  |  | CPE + replicates / Total replicates |  |  |  |  |
| --- | --- | --- | --- | --- | --- | --- |
| Sample Collection Method | Sample | Mock | 1% Triton X-100 Treatment Time |  |  |  |
|  |  |  | 0 minutes | 30 minutes | 1 hour | 2 hours |
| Salivette | 1 | 0/2 | 3/3 | 0/3 | 0/3 | 0/3 |
|  | 2 | 0/2 | 3/3 | 0/3 | 0/3 | 0/3 |
|  | 3 | 0/2 | 3/3 | 0/3 | 0/3 | 0/3 |
|  | 4 | 0/2 | 3/3 | 0/3 | 0/3 | 0/3 |
|  | 5 | 0/2 | 3/3 | 0/3 | 0/3 | 0/3 |
|  | 6 | 0/2 | 3/3 | 0/3 | 0/3 | 0/3 |
| Saliva into Tube | 1 | 0/2 | 3/3 | 3/3** | 0/3 | 0/3 |
|  | 2 | 0/2 | 3/3 | 0/3 | 0/3 | 0/3 |
|  | 3 | 0/2 | 3/3 | 0/3 | 0/3 | 0/3 |
|  | 4 | 0/2 | 3/3 | 2/3** | 0/3 | 0/3 |
|  | 5 | 0/2 | 3/3 | 0/3 | 0/3 | 0/3 |
|  | 6 | 0/2 | 3/3 | 0/3 | 0/3 | 0/3 |

\* Cytopathic Effect (CPE) observed in Vero E6 cells 5 days post-inoculation with saliva samples spiked with SARS-CoV-2 and subsequently treated with 1% Triton X-100 for varying times. Mock replicates were inoculated with saliva + media. All other replicates contained saliva + virus + Triton X-100 for the indicated time. Red cells indicate SARS-CoV-2 non-inactivating conditions. Green cells indicate SARS-CoV-2 inactivating conditions.
\*\*CPE+ wells did not show the same effect upon passage.

**Table S3.** ROC statistics table for ELISAs conducted on saliva samples*.

|  | *Manual IgG* |  | *Manual IgA* |  | *Manual IgM* |  |
| --- | --- | --- | --- | --- | --- | --- |
| Marker | Spike | RBD | Spike | RBD | Spike | RBD |
| AUC | 0.970 | 0.947 | 0.837 | 0.764 | 0.866 | 0.793 |
| SE.AUC | 0.0139 | 0.0187 | 0.0341 | 0.0412 | 0.0328 | 0.0394 |
| Lower Limit | 0.943 | 0.911 | 0.770 | 0.683 | 0.801 | 0.716 |
| Upper Limit | 0.998 | 0.984 | 0.903 | 0.845 | 0.930 | 0.870 |
| z | 33.77 | 23.900 | 9.872 | 6.399 | 11.146 | 7.434 |
| p-value | 0 | 0 | 0 | 0 | 0 | 0 |
| *ROC Coordinate points at $\leq 2\%$ FDR* |  |  |  |  |  |  |
| Marker | Spike | RBD | Spike | RBD | Spike | RBD |
| Cutpoint | 5.550 | 3.341 | 12.172 | 42.580 | 2.564 | 3.291 |
| FPR | 0.0196 | 0.0196 | 0.0196 | 0.0196 | 0.02 | 0.02 |
| TPR | 0.886 | 0.848 | 0.506 | 0.304 | 0.571 | 0.325 |
| *ROC Coordinate points at $\leq 4\%$ FDR* |  |  |  |  |  |  |
| Marker | Spike | RBD | Spike | RBD | Spike | RBD |
| Cutpoint | 2.969 | 2.283 | 8.667 | 27.270 | 1.428 | 2.595 |
| FPR | 0.0392 | 0.0392 | 0.0392 | 0.0392 | 0.04 | 0.04 |
| TPR | 0.924 | 0.848 | 0.608 | 0.430 | 0.688 | 0.390 |
| *Samples used to generate ROC curves* |  |  |  |  |  |  |
| Sample Cohort | No. Samples |  |  |  | No. Samples |  |
| COVID-19 Patient Samples | 79 |  |  |  | 77 |  |
| Pre-COVID-19 Negative Controls | 27 |  |  |  | 27 |  |
| Unexposed Negative Controls Collected in 2020 | 24 |  |  |  | 23 |  |

\* Sensitivity values reported in the manuscript are based on the coordinate points at a false positive rate of $\leq 2\%$

